# A Methodological Evaluation of Meta-Analyses in tDCS - Motor Learning Research

**DOI:** 10.1101/2024.07.26.24311068

**Authors:** Taym Alsalti, Ian Hussey, Malte Elson, Robert Krause, Steffi Pohl

## Abstract

With transcranial direct-current stimulation’s (tDCS) rising popularity both in motor learning research and as a commercial product, it is becoming increasingly important that the quality of evidence on its effectiveness be evaluated. Special attention should be paid to meta-analyses, as they usually have a large impact on research and clinical practice. The aim of this study was to evaluate the methodological quality of meta-analyses estimating the effect of tDCS on motor learning with respect to reproducibility as the main focus, and reporting quality and publication bias control as secondary aspects. The three meta-analyses we reviewed largely adhered to PRISMA reporting guidelines and reported the primary effect sizes and sampling variances / confidence intervals they calculated, enabling successful reproductions of pooled effect size estimates. However, akin to previous meta-research reviews with similar aims, we found the methods and results sections of the meta-analyses to be severely underreported, which compromises the ability to judge the soundness of the methodological procedure adopted as well as its reproducibility. While publication bias detection methods were applied, the approaches chosen do not allow for well informed decisions about the presence or extent of publication bias. These results reemphasise the need to transparently report methods in meta-analyses and to meticulously evaluate their quality before and after publication.

## Introduction

### tDCS and its applications

Transcranial direct-current stimulation (tDCS) is a non-invasive brain stimulation technique which involves delivering constant, low current to the brain via electrodes fixed on the scalp (Gazzaniga et al., 2018). tDCS is believed to be safe and to cause negligible side-effects, if any (Gianni et al., 2021; Nitsche et al., 2008). The typical set-up of a tDCS protocol involves no more than a handful of inexpensive components and relatively simple steps (Gebodh et al., 2019; Woods et al., 2016). Experimental paradigms aiming to evaluate such tDCS effect typically take the following form: after baseline measurements of different outcomes on the first day, study participants undergo a series of training sessions in a specific motor task (e.g., squeezing a hand-held force transducer to move the cursor on the computer screen in a certain manner). The participants receive either tDCS stimulation or get the device placed on their heads without administering any current (sham/control condition) during this training. Performance is measured subsequently (online) and/or 1 to several weeks or months later (consolidation/retention).

When considering tDCS’ logistic advantages, tDCS’s rapidly increasing popularity over the last two decades is not surprising (Buch et al., 2017). In clinical research, its efficacy for treating or attenuating depression (Brunoni et al., 2016), memory deficits in Alzheimer’s patients (Bennabi et al., 2014), pain (Luedtke et al., 2012), schizophrenia (Liu et al., 2021), among other conditions, has been investigated. Interest in tDCS is not restricted to clinical settings: studies on healthy subjects have been conducted to test its effects on cognitive abilities such as language and memory (Horvath et al., 2015), affective states (Austin et al., 2016), and motor skills, such as surgery (Hung et al., 2021) and musical performance (Rosen et al., 2016).

tDCS has become so established as a research tool that it is already approved for clinical use (with some restrictions) in several countries around the world (Fregni et al., 2015). Furthermore, an ever-richer variety of commercial tDCS products has become available on the market (Davis, 2016; Wexler, 2018; Zettler, 2017), prompting experts (e.g., Wurzman et al., 2016) to voice concerns over the increasing prevalence of this “do-it-yourself” use of tDCS. Out of the 449 such at-home tDCS consumers surveyed by (2018), 52% (237) did so for enhancement purposes. tDCS devices have also become available as commercial products in many countries (e.g., this device by Neurosym that advertises “Vagus Nerve Stimulation for a Better Health”). The touted benefits range from alleviating depressive symptoms to improving physical strength and dexterity. At the same time, heterogeneity in technical implementations (e.g., simulation intensity and duration) and problems like publication bias and irreproducibility plague the tDCS literature (Buch et al., 2017). This also impacts meta-analyses on this topic, which often form the basis of clinical decisions.

### Methodological quality of Meta-analyses

The popularity of tDCS might be partially driven by meta-analyses showing positive effects of its use. Meta-analyses are often characterised as being at the “top” of the “evidence hierarchy” (Evans, 2003): they are cited more frequently than primary studies about the same topics, are commonly assumed to provide the most accurate estimate of an effect, and have a large impact on theory development as well as policy and clinical practice (Gopalakrishnan & Ganeshkumar, 2013; Gøtzsche et al., 2007; Ioannidis, 2016; Lakens et al., 2017; Morganti, 2007). Several papers drafting guidelines on the use of tDCS in research and the clinic extensively cite meta-analyses (Charvet et al., 2020; Fregni et al., 2015; Lefaucheur, 2016). Hence, meta-analyses on the effects of tDCS are likely to influence its uptake in many areas. Given this, it is important to evaluate the methodological quality of these meta-analyses as they can inform clinical practice. There are different aspects of methodological quality that can be considered, such as comprehensiveness of the study search, quality of the study screening procedure, and appropriateness of statistical methods employed. Here, we focus on reporting quality, reproducibility, and publication bias control.

#### Reporting quality of meta-analyses

Meta-analysts are faced with a plethora of different decisions with regards to which databases to search and using which strings; primary study selection and exclusion; data extraction; statistical methods, among others (Ada et al., 2012; Guzzo et al., 1987; Valentine et al., 2010; Voracek et al., 2019). Although the extent to which these “researcher degrees of freedom” (Simmons et al., 2011) impact the conclusions of the meta-analysis is not a completely uncontroversial issue^1^, there is consensus regarding the importance of transparently reporting these decisions (Aguinis, Pierce, et al., 2011), as it is difficult to assess the quality and trustworthiness of that which one does not have access to (Page, McKenzie, et al., 2021).

Several guidelines for conducting and reporting meta-analyses exist, e.g., the Cochrane Handbook (a comprehensive guide for conducting systematic reviews and meta-analyses, Higgins et al., 2019) or the Preferred Reporting Items for Systematic Reviews and Meta-Analyses (PRISMA, a checklist of methodology reported items to report when conducting systematic reviews and meta-analyses Moher et al., 2000, 2009; Page, McKenzie, et al., 2021). Although such guidelines have been available for over two decades, meta-scientific evaluations of adherence to PRISMA and other guidelines have shown that reporting standards of meta-analyses are generally suboptimal and that these guidelines are rarely fully adhered to (Page & Moher, 2017; Polanin et al., 2020; Schalken & Rietbergen, 2017). For example, while over 80% of the +80000 systematic reviews coded by Page & Moher (2017) provided a rationale for their review, less than 60% conducted a risk of bias analysis or disclosed funding sources.

#### Meta-analysis reproducibility

Given the prevalent lack of adherence to reporting checklists, it comes as no surprise that attempts to reproduce meta-analyses often fail; the less information a meta-analyst provides about their procedure, the harder it is to reproduce (Aguinis, Pierce, et al., 2011). In fact, even full adherence to reporting guidelines far from guarantees reproducibility (Weissgerber et al., 2021). Reproducibility is essential as it enables other researchers to detect errors and evaluate the defensibility of subjective choices (e.g., primary study eligibility criteria). It also facilitates updating meta-analyses as more relevant primary studies become available (Lakens et al., 2016). Reviews of meta-analysis reproducibility (e.g., Ford et al., 2010; Gøtzsche et al., 2007; Lakens et al., 2017; Maassen et al., 2020) emphasised somewhat different methodological aspects (e.g., complete reporting vs. computational correctness) but mostly focused on reproducing data extraction and effect size (ES) computation (both primary and pooled). Their conclusions about the reproducibility of meta-analyses were, although of varyingly grave consequences^2^, also similar: the reproducibility of meta-analyses was severely limited due to under-reporting and errors.

#### Publication bias control

Publication bias, that is, the tendency of researchers to suppress findings that do not go their way and journals to selectively publish studies that report significant results, remains a major concern. Meta-analysts can attempt to both pre-emptively mitigate publication bias, e.g., by searching the “grey literature” for unpublished studies, and through the post-hoc application of statistical publication bias detection and adjustment methods. Many procedures for this latter purpose have been developed in the last three decades, displaying varying performance profiles depending on assumptions made and nature of data (see e.g., Harrer et al., 2019 for a detailed review). Besides agreement that traditional methods like Fail-Safe *N* and Trim-and-Fill are limitedly informative, there is little agreement about which specific methods should be used under which conditions (Carter et al., 2019; McShane et al., 2016; Renkewitz & Keiner, 2019). This necessitates transparently discussing the assumption of each test used and to what extent these assumptions are met in the current set of primary studies. Another commonly voiced recommendation is to use several methods in tandem as sensitivity analyses, and not as detection or adjustment methods per se (e.g., Vevea et al., 2019).

### Research goals

In sum, four principal premises motivated this work: 1. tDCS appears to be remarkably popular as a research tool in basic and clinical research as well as in form of commercial gadgets, 2. meta-analyses of tDCS’s effect on motor learning might be substantially impacting research and clinical practice and, further downstream, tDCS’s uptake as a commercial product, 3. The credibility and informativeness of meta-analyses depend on their methodological quality, 4. no methodological evaluation of meta-analyses in tDCS-motor learning research has been conducted as of yet. Our aim was thus to evaluate the methodological quality of meta-analyses in the tDCS motor learning literature with respect to (1) reproducibility (primary aim) as well as (2) adherence to PRISMA guidelines and (3) evaluation of and controlling for publication bias (secondary aims).

## Methods

Although our methodological approach mostly followed the plan pre-defined in the thesis proposal (accessible on the thesis’ Open Science Framework [OSF] project osf.io/xyhf5), there were important deviations from the plan, especially with respect to reproducibility testing. A document listing these deviations and reasons for them can be found on the OSF project (osf.io/8w4v6). For data wrangling, analysis, and visualisation, we used R (R Core Team, 2021) and the packages dmetar (Harrer et al., 2019), dplyr (Wickham et al., 2021), ggplot2 (Wickham, 2016), MAd (Hoyt, 2014), Matrix (Bates & Maechler, 2021), meta (Balduzzi et al., 2019), metafor (Viechtbauer, 2010), purrr (Henry & Wickham, 2020), and tidyr (Wickham, 2021). The package groundhog (https://cran.r-project.org/package=groundhog) was used to ensure long-term reproducibility of our analyses. Data were extracted from figures using WebPlotDigitizer (https://wpd.starrydata2.org/, Rohatgi, 2021). A video demonstration of how we extracted data from figures is available on our OSF project. All our data and code are available on the project’s GitHub repository, github.com/TaymAlsalti/tDCS_meta-analysis.

### Sample of meta-analyses

We selected three English-language meta-analyses (Hung et al., 2021; Kang et al., 2016, 2018) based on the following eligibility criteria:

- Meta-analysis studies which quantitatively synthesise multiple (at least 3) primary studies on the effects of tDCS on motor learning.
- No restriction on primary outcomes (e.g., how speed or accuracy were measured), designs of primary studies (e.g., randomised vs. crossover designs), or participants (e.g., clinical or healthy subjects) in the primary studies were imposed.

Exclusion criteria:

- Reviews of any type without a quantitative synthesis
- Primary studies
- Reviews which did not report a “main” meta-analysis, but rather multiple meta-analyses of subgroups of studies.

### Reproducibility

We based our treatment of meta-analysis reproducibility on the definition and principles of reproducibility put forward by the American Statistical Association (Broman et al., 2017): a meta-analysis is reproducible if its authors provided enough information to go through all the necessary procedures (search, screening, data extraction, calculation of primary ESs, calculation of the pooled ES…) to arrive at the same numerical results. However, although we acknowledge the importance of all these steps, we, like Gøtzsche et al. (2007), Lakens et al. (2017), and Maassen et al. (2020), focused on data extraction and calculation of ES estimates.

For each meta-analysis selected, we extracted information on both the primary and meta-analyses study levels. On the primary study level, we extracted all information necessary to calculate an ES measure as well as its standard error (SE). This included information such as sample sizes, means and SDs of outcome measures in the compared groups. We extracted these data from both the primary studies and the meta-analyses, whenever reported. On the meta-analysis level, we additionally extracted the pooled ES estimate across the primary studies.

*Reproducing primary ESs*. In order to reproduce the pooled ES from the meta-analyses, it is necessary to reproduce the ESs from the primary studies (primary ESs) first^3^. For classifying the reproducibility status of the primary ESs, we constructed a scheme with two variables (see Table 1): A. whether the primary ES could be successfully reproduced numerically (results reproducibility, Goodman et al., 2016). Here, an ES was considered successfully reproduced (or reproducible) if the reproduced ES equalled the one reported in the meta-analysis at the second decimal (e.g., 0.3334543 = 0.33). We considered an ES to be approximated (but not reproduced) if the reproduced ES values were in a ±0.05 range of the reported values in the meta-analysis^4^. And B. whether the procedure we followed in reproducing the ES strictly corresponded to the information given in the meta-analysis or to the procedure apparently adopted for at least two other ES*sp*included in the meta-analysis (methods reproducibility). “Procedure” here includes such analytic decisions as using *p*-values or test statistics in combination with sample sizes, using the raw means and SDs, using means and SDs of changes in the outcome from baseline, etc. to estimate a standardized mean difference (ES) for a given primary study. The second variable in the classification system was thus mainly adopted to capture cases where there was a discrepancy between how the meta-analysts reported having computed a primary ES and how they actually computed it.

**Table 1.**
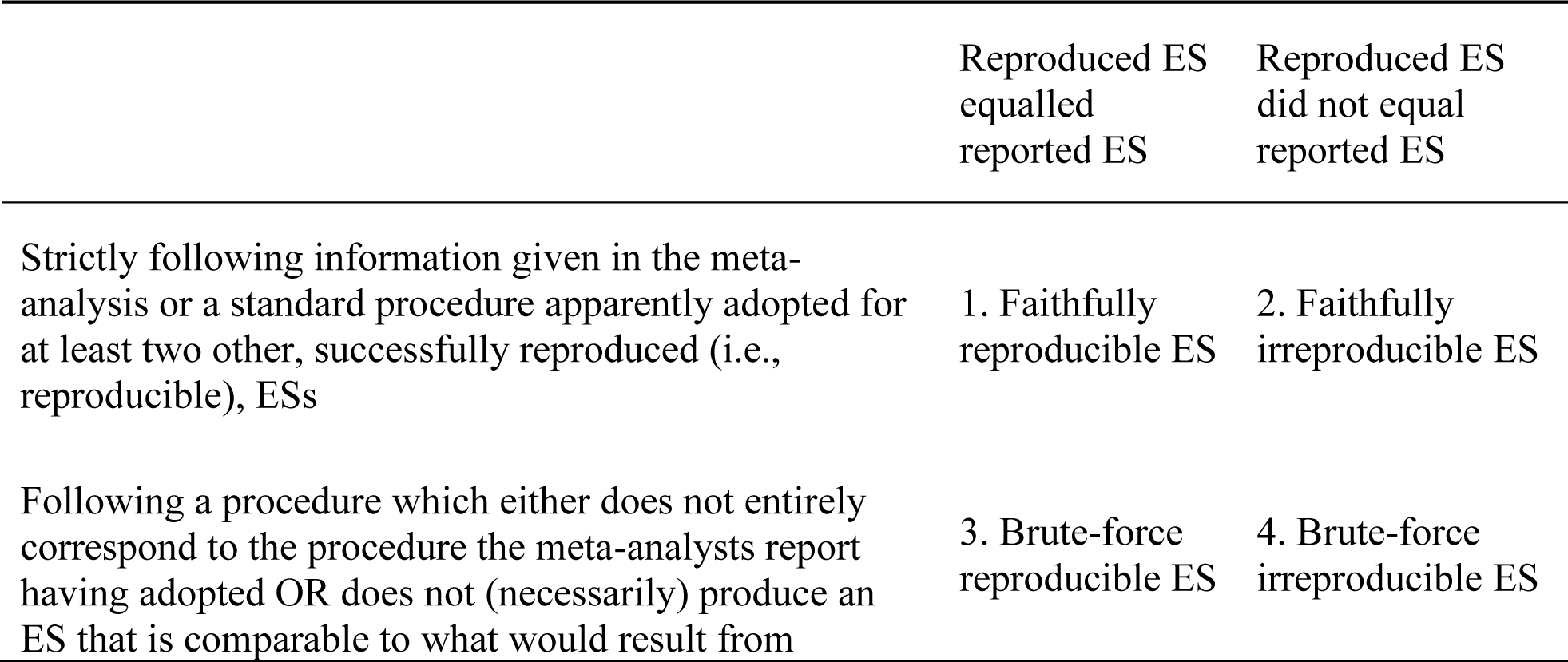
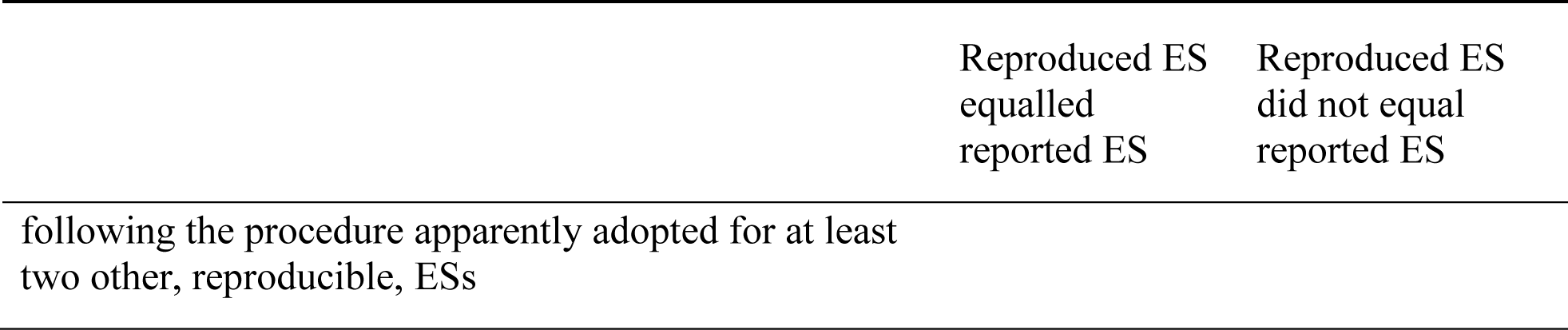
Reproducibility classification scheme for primary effect sizes.

|  | Reproduced ES<br>equalled<br>reported ES | Reproduced ES<br>did not equal<br>reported ES |
| --- | --- | --- |
| Strictly following information given in the meta-analysis or a standard procedure apparently adopted for at least two other, successfully reproduced (i.e., reproducible), ESs | 1. Faithfully<br>reproducible ES | 2. Faithfully<br>irreproducible ES |
| Following a procedure which either does not entirely correspond to the procedure the meta-analysts report having adopted OR does not (necessarily) produce an ES that is comparable to what would result from | 3. Brute-force<br>reproducible ES | 4. Brute-force<br>irreproducible ES |
| following the procedure apparently adopted for at least<br>two other, reproducible, ESs |  |  |

The distinction between “reproduced” and “reproducible” here is thus crucial: “reproduced” means that we calculated an ES based on data extracted from the primary study that we believed the meta-analysts might have used, whereas “reproducible” primary ESs were those which numerically equalled the ones reported in the meta-analysis. Whereas “faithfully” and “brute-force” refer to whether the meta-analysis provided enough information on how to compute the primary ES. It follows from this scheme that primary ESs that cannot be reproduced due to lack of information cannot be tested for numerical reproducibility, and thus cannot be faithfully reproducible, although they can be brute-force reproducible. Also note that if we classify an ES as “faithfully irreproducible”, this does not imply that it is necessarily also brute-force irreproducible because in most cases, we did not collect further values to test brute-force reproducibility if the values we chose to test faithful reproducibility very clearly corresponded to the meta-analysts’ description of their procedure.

Reproducibility testing was an iterative process which involved several rounds of data extraction. The initial round of data extraction and testing reproducibility yielded few reproducible ESs as most primary studies did not report the values necessary to directly compute an ES and/or its sampling variance (e.g., for a between groups Cohen’s *d* this would be the group means, SDs and sample sizes). Therefore, a less strict data extraction procedure was adopted, which involved the following steps:

1. For each primary ES, we first looked for the raw means and SDs of the outcome reported as having been used by the meta-analysts. If the primary study reported multiple sets of means and SDs which can be seen as corresponding to the outcome described in the meta-analysis (e.g., the outcome in the meta-analysis for a given primary study is “Fugl-Myer Test” but the primary study reports values for “Fugl-Myer Test-upper limbs” and “Fugl-Myer Test-full”), all sets were extracted.
2. If no means and SDs for the relevant outcome were reported in the primary study, means and SDs were extracted from figures. If no figures were reported which contained means and SDs (or SEs or confidence intervals [CIs], which can be converted to SDs), *p* and/or *t*-values for tests on the relevant outcome were extracted, which in combination with sample sizes can be converted to ESs.
3. Based on all extracted values, we computed each primary ES using the estimator (Cohen’s *d* or Hedges’ *g*) reported as having been used by the meta-analysts. If this information was not given in the meta-analysis, we tried both formulas and for further analysis used the one which consistently approximated the reported ESs better.
4. If none of the values extracted reproduced a given ES, we double checked the correctness of the data extracted and, in some cases, extracted more values from the primary study (à la brute-force) and computed the ES based on those.
5. We computed the sampling variances based on the ESs and the corresponding samples sizes.

The concrete procedure for data extraction thus differed for each single primary study. A detailed description of all values we extracted and how we analysed them is provided in the data analysis notebook, also available on our OSF project page.

#### Reproducing the pooled ESs

For obtaining an estimate of the pooled ES, we conducted three analyses based on different subsets of primary ES: (a) using the primary ESs and sampling variance as reported in the meta-analysis^5^, (b) using the faithfully reproduced and faithfully approximated in addition to the brute-force reproduced primary ESs, and (c) using only the faithfully reproduced primary ESs. This allowed us to investigate to which extent differences between reported and reproduced pooled ESs are due to problems to reproduce primary ESs and which are due to issues with the description of the meta-analytic procedure.

### Adherence to PRISMA reporting guidelines

We evaluated adherence to PRISMA reporting guidelines (Liberati et al., 2009; Moher et al., 2009) as they are the most widely adopted reporting standard for meta-analysis. To that end, we coded whether the meta-analysis reported the relevant information as recommended for each of the 27 items, regardless of whether the meta-analysis reported having adhered to any reporting guidelines. Using the PRISMA checklist, we scanned the full text of each meta-analysis, searched for keywords pertaining to each item (e.g., “search” for the item “Search”) and noted down the item as reported regardless of how well it was described.

### Publication bias

We investigated in which way the meta-analyses in our study considered the possible impact of publication bias by (a) evaluating measures taken to identify unpublished studies and (b) assessing whether and which statistical approaches were used to investigate the presence of publication bias. We additionally coded whether the meta-analysts took measures to:

1. Searched clinical trial registries (e.g., ClinicalTrials.org)
2. Searched thesis and dissertation repositories (e.g., ProQuest)
3. Contacted known researchers in the field to inquire about unpublished results
4. Contacted authors of included studies to ask for raw data or unpublished results Besides coding whether any statistical methods were used at all and which, we additionally tested for publication bias in each meta-analysis using 3 different publication bias methods not used by the meta-analysts to evaluate the robustness of their conclusions (see Supplementary Note 1).

### Additional coding/analyses

We checked whether the meta-analyses provided data-analysis code, shared their extracted data, or had been pre-registered. As outliers can heavily impact pooled ES estimates, we coded whether and how the meta-analyses tested for the existence of outliers among the included studies and, based on the data extracted from the forest plots in the meta-analyses, we tested for the impact of outliers on the results ourselves using the leave-one-out method (Harrer et al., 2021; Tobias, 1999).

## Results

### Sample of meta-analyses

We selected the first three meta-analyses we found via non-systematic Google Scholar and Web of Science searches and which fulfilled our eligibility criteria. Although we later also identified other meta-analyses, which based on the abstract might have been eligible, we did not screen full texts to check eligibility for feasibility reasons. In that sense, our sample of three meta-analyses was a “convenience” sample. Table 2 gives an overview of the three meta-analyses selected. Meta-analyses 1 and 2 aimed to estimate the effectiveness of tDCS for improving motor function in post-stroke patients, although meta-analysis 2 focused exclusively on the effects of cathodal tDCS. Meta-analysis 3 investigated effectiveness of tDCS for improving surgical performance of surgery trainees. The first meta-analysis synthesised the results of 13 randomised controlled trials (RCTs) and 4 crossover trials, the second 6 RCTs and 9 crossover trials, the third 5 RCTs and one crossover trial.

**Table 2.**
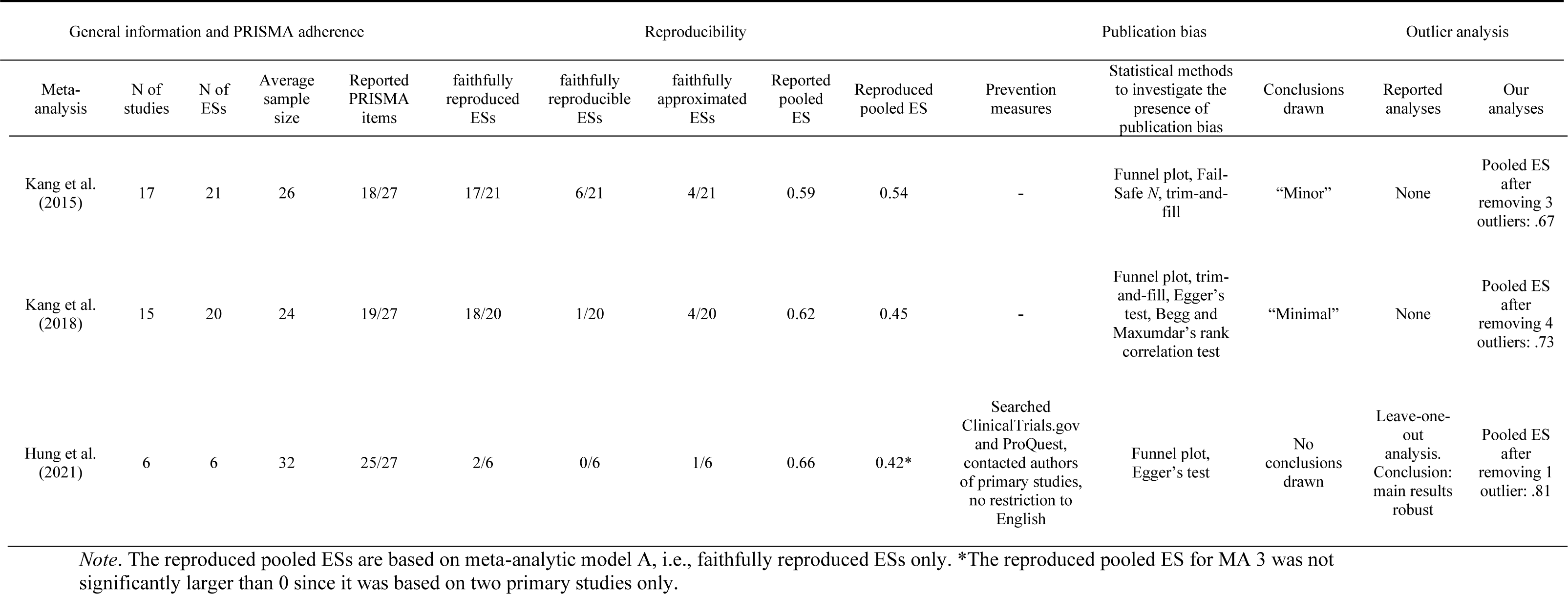
Reviewed meta-analyses.

All three meta-analyses reported ESs in form of standardised mean differences (SMDs), although it was not consistent which group means were used (e.g., mean difference between pre and post vs. mean difference in the post values across groups). For some primary studies, meta-analyses 1 and 2 included more than one ES into the meta-analysis. In meta-analysis 1, these represented cathodal vs. sham and anodal vs. sham pairs, whereas in meta-analysis 2, the different ESs within one study were based on two different outcomes. None of the three meta-analyses had been pre-registered, shared data, or provided data analysis code. Meta-analysis 3 stated that data “was available upon reasonable request” (p. 11), but the corresponding author of the meta-analysis did not respond to our email requesting more information/data.

### Reproducibility

*Reproducibility of primary ESs*. Most primary ESs in meta-analyses 1 and 2 (∼81% and ∼85%, respectively) and 33% in MA 3 could be faithfully reproduced (see Table 2). That is, (a) enough information was available or inferable from the meta-analyses and (b) seemingly appropriate data were reported in the corresponding primary studies, to attempt to recalculate about 77% of all primary studies across the 3 meta-analyses. Of those, ∼58%, 27%, and 50%, respectively, were reproducible (i.e., successfully reproduced numerically) or approximated.

We faced considerable difficulties in reproducing the primary ESs for all three meta-analyses, mainly due to limited reporting of their methods sections. The process necessitated several rounds of data extraction and testing. It was in most cases impossible to know from the paper how the meta-analysts calculated each primary ES and in a brute-force procedure, we had to rely on trial and error to figure out which values from the primary studies were used. Table 3 lists the relevant pieces of information which were reported or missing from the three meta-analyses. As can be seen from Table 3, while sample sizes and groups being compared were reported by all three meta-analyse, none of them reported enough information on the outcome and exact calculation method to ensure unambiguity.

**Table 3.**
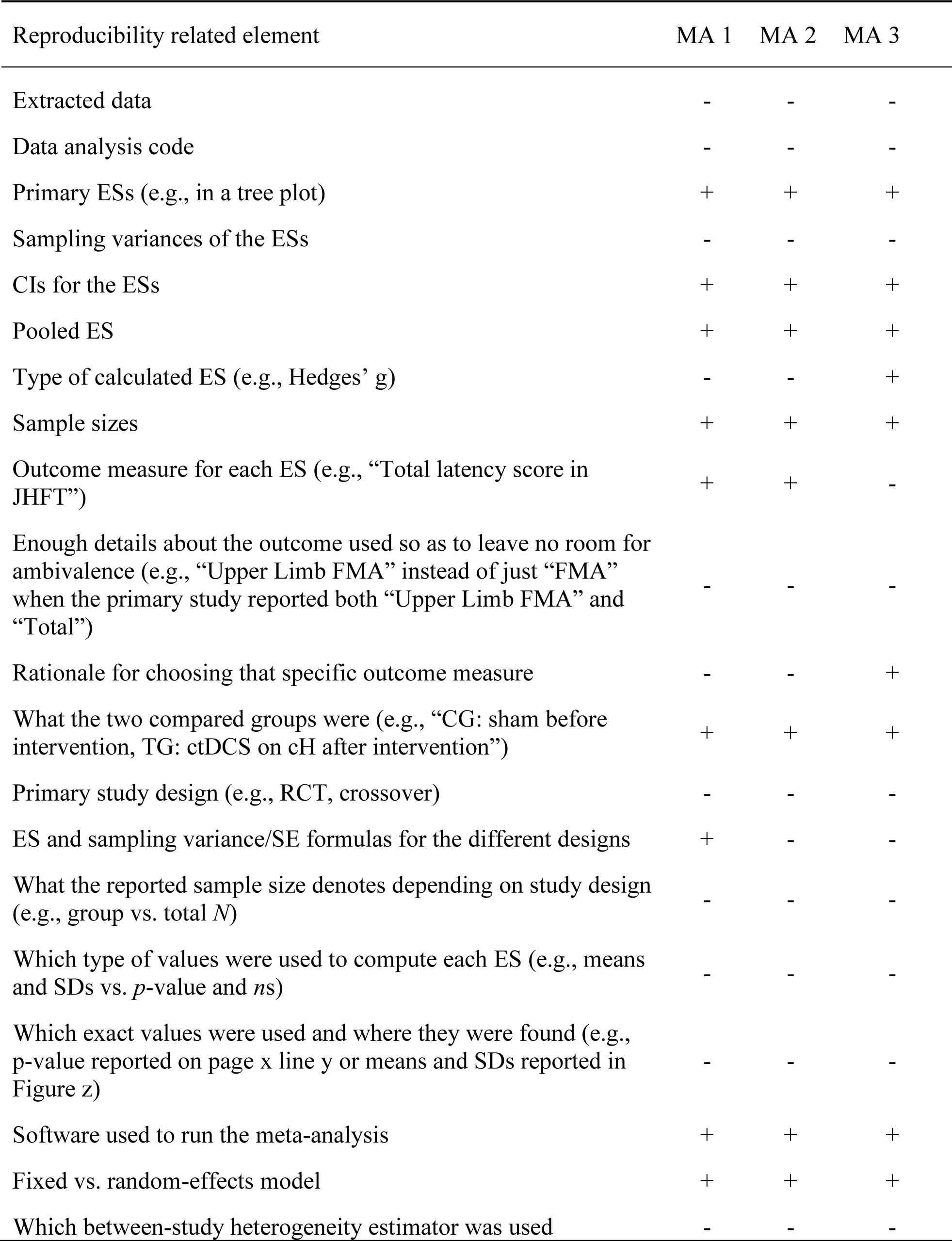
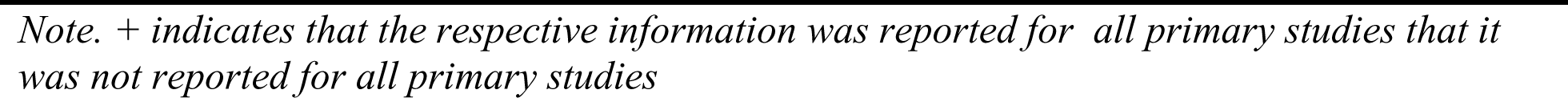
Reproducibility related elements and whether each meta-analysis (MA) provided them.

In most cases, it was unclear why we failed to reproduce any given primary ES, but our extensive attempts at brute-force reproducing certain primary ESs which ended up exactly reproducing reported ESs gave us some insights for reasons why we could not faithfully reproduce these ESs: For example, all three meta-analyses used values belonging to different outcomes than specified and used *p*-values from nonparametric tests (e.g., median tests). The third MA used *p*-values expressed as a range (e.g., *p* <.01). Tables S1-3 list all reproduced-reported primary ES pairs for all three meta-analyses, their corresponding reproducibility classification, and the potential reason for irreproducibility, if applicable. Figure 1 depicts all primary ESs reported in the forest plots shown in three meta-analyses and how they compared to their reproduced counterparts. Across all 3 meta-analyses, reproduced ESs were on average.18 smaller than reported ones (max = 1.08, min = −2.99). The average absolute difference was.37 (max = 2.99, min = 0).

**Figure 1:**
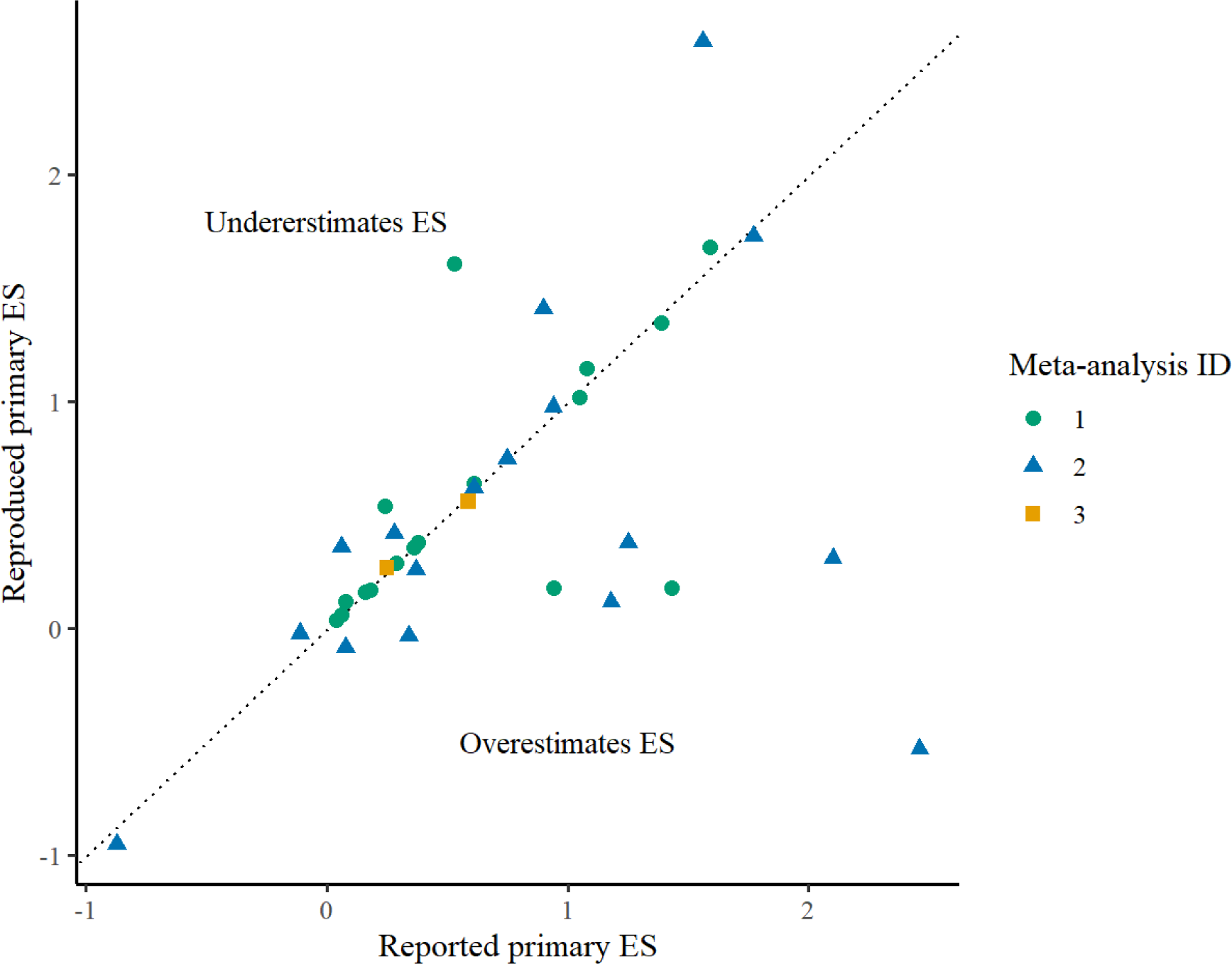
Comparison of ESs reported in the three meta-analyses and their faithfully reproduced counterparts. The 13 ESs that could not be faithfully reproduced are not depicted (see Figure S1 for an equivalent plot showing both faithfully and brute-force reproduced ESs).

*Reproducibility of pooled ESs*. Since all three meta-analyses reported having fit a random-effects model using the Comprehensive Meta-Analysis software^6^, which per default estimates between-study heterogeneity via the Der-Simonian-Laird method (DerSimonian & Laird, 1986), we used these settings for all our analyses, too. Whereas the pooled ESs we calculated using the primary ESs reported in the meta-analyses and the sampling variances extracted from funnel plots or converted from CIs were reproducible, those based on faithfully reproduced primary ESs were on average.15 points smaller than the reported ones. The largest impact was suffered by the already small MA3, whose pooled ES was not significant based on the two faithfully reproduced ESs. When adding the brute-force reproduced ESs to the faithfully reproduced ones, we got pooled ESs of.70,.40,.65, respectively, for the three meta-analyses.

### PRISMA adherence

All three meta-analyses reported most of the items given in the PRISMA guidelines. Meta-analyses 1 and 2 did not report having adhered to any reporting guidelines. Despite this, they can be seen as having reported the content of 18 and 19 items, respectively, out of the 27 PRISMA (Moher et al., 2009) items. Meta-analysis 3 reported having adhered to the most recent PRISMA guidelines (Page, McKenzie, et al., 2021) but we evaluated the adherence to the items of the 2009 version to ensure comparability to the other meta-analyses. Meta-analysis 3 reported the content of 25 out of the 27 items. To highlight one aspect, all three meta-analyses adequately described their eligibility criteria but none of them described the actual process of how the criteria were enforced.

### Publication bias

Table 2 gives an overview of measures taken by the meta-analysts to investigate or control for publication bias. All three meta-analyses mentioned publication bias and applied a number of different tests to investigate the presence of publication bias. They either concluded that there is hardly any bias or did not draw a conclusion at all. The authors of meta-analysis 1 concluded that the findings of the tests they used “support a minor publication bias conclusion” (Kang et al., 2016, p. 348). Similarly, the conclusion in meta-analysis 2 (Kang et al., 2018, p. 5) was “minimal publication bias in the studies used”. No clear conclusion was provided in meta-analysis 3. Of all three meta-analyses, only meta-analysis 3 reported having taken measures to pre-emptively mitigate its effects. These included a number of different measures aiming at identifying unpublished studies.

### Additional coding/analyses

Only the third meta-analysis mentioned outliers or influential studies. They ran a leave-one-out analysis and reported that the main results did not change due to removing any one of the 6 studies they included. Our leave-one-out analysis indicated the presence of influential studies in all three meta-analyses (see Figures S2-S4). Attempting to reproduce the meta-analyses revealed further methodological issues which do not directly pertain to the main methodological aspects we investigated: All three meta-analyses indiscriminately combined primary studies of different designs.

For example, they computed ESs using the same formula for both controlled and crossover designs, a procedure which neglects bias resulting from estimating sampling variances for crossover studies without accounting for carry-over effects or correlations between time points (Borenstein & Hedges, 2019; Madeyski & Kitchenham, 2018; Morris & DeShon, 2002). Furthermore, the fact the meta-analysts used the total sample size of the cross-over trial to replace the treatment and control group sample sizes is likely to have inflated the power of these pair-wise comparisons (which is especially relevant for the publication bias tests). Another issue the meta-analysts neglected to account for is ES dependency (Gleser & Olkin, 2009), which is especially critical in the case of the first two meta-analyses as they derived multiple ESs from single studies.

## Discussion

The aim of this work was to evaluate the methodological quality of meta-analyses in tDCS-motor learning research with respect to reporting quality, reproducibility, and publication bias control. We found that although the meta-analyses largely fulfilled reporting requirements like PRISMA (Moher et al., 2009), they were too underreported to allow for smooth reproductions. While pooled ESs were reproducible based on values extracted from tables or figures (and default software settings for heterogeneity estimators), a considerable number of primary ESs was not. The reported primary ESs were on average larger than the ones reproduced. Consequently, we failed to numerically reproduce the pooled ES estimates reported in all meta-analyses when following the procedures they described. As to publication bias control, only meta-analysis 3 reported having searched the grey literature. While all three meta-analyses used several publication-bias detection methods, they mostly used “traditional” approaches without discussion of their assumptions or appropriateness.

The most notable finding is probably the high prevalence of discrepancies between how the meta-analysts reported having computed individual ESs and how they apparently did it. These discrepancies were most often in relation to the outcomes used. For example, there were multiple cases where the meta-analysis reported having used an outcome X whereas they seemed to have used the outcome change in X from baseline. This was particularly perplexing when values for both outcome and outcome change were reported in the primary study. In general, all primary studies included in the meta-analyses reported values/tests for several outcomes and meta-analysis 3 was the only one to provide a rationale, albeit a vague one^7^, for why they chose the outcome they did for each primary study. In most cases, it was impossible to infer how these things came about as the authors of meta-analyses 1 and 2 did not respond to our request for data or data analysis code or protocol and the authors of meta-analysis 3 did not respond to our email at all. On the whole, we had similar difficulties in our reproducibility endeavour as Lakens et al. (2017) and Maassen et al. (2020).

Although our reproduced pooled ESs were calculated using less primary ESs for all three meta-analyses (due to methodologically irreproducible ESs), none of our reproductions led to a radical change in the pooled ES estimate like in Gøtzsche et al.’s (2007) or Ford et al.’s (2010) reviews. For example, none of the pooled ES estimates was completely nullified or changed its sign in the reproduced version. However, we documented several haphazard ways some primary ESs were calculated as well as some errors. Given this, a reproduction that is compliant with methodological guidelines rather than attempting to be faithful to the meta-analysts’ workflow might very well have led to different results. There are some indications that the meta-analysts’ deviations from their procedure led to overestimating the pooled ESs.

### Recommendations

Although the meta-analyses we reviewed were adequate in some respects (e.g., they all reported a forest plot displaying all ESs and their CIs), there is clearly some room for improvement. As a remedy for the compromised reporting quality and reproducibility, more detailed descriptions of the methodological procedure are called for, ideally accompanied by raw data and the data analysis code (Lakens et al., 2016; Page, Moher, et al., 2021). As publication bias may heavily impact the results of meta-analyses, we recommend to a) take measures to control for publication bias (Vevea et al., 2019) and b) apply a more extensive testing procedure involving sensitivity analyses using different methods and different parameter settings within a method and discussion of the respective assumptions. When dealing with a highly heterogeneous set of ESs, van Aert et al. (2016) recommend splitting the set into subgroups based on theoretical and methodological considerations before carrying out the bias testing procedure. Inzlicht et al. (2015) recommend presenting a range of plausible pooled ESs based on different publication bias tests (using different assumptions).

No self-evident solutions can be offered for the shortcomings related to the synthesis-related aspects of the meta-analyses. There is immense variation in reporting quality of primary studies and meta-analysts usually have no other choice but to work with the data reported in the primary study (that is, when contacting the authors of the primary study for more data is not feasible or proves to be fruitless, Higgins et al., 2019). It is thus entirely understandable that meta-analysts must sometimes resort to alternative means of calculating certain measures. However, there are better ways to do this, too, than what the meta-analysts seemingly opted to do. For example, instead of converting a *p*-value derived from a medians test to an ES, the meta-analysts could have estimated the means and SDs based on the medians and the corresponding interquartile ranges and calculated the ES based on these means and SDs, which would have yielded a more comparable result to an ES calculated based on actual means and SDs (Hozo et al., 2005; Wan et al., 2014). Similarly, there are methods to compute the ESs and their corresponding sampling variances when dealing with crossover studies (Madeyski & Kitchenham, 2018). Estimating the ESs using the formula for within-subjects designs would have taken the correlations between time points into account (Borenstein & Hedges, 2019).

### Limitations

There are several limitations of our study. First, we defined our exclusion criteria based on largely practical considerations. Their high restrictiveness has probably led to a sample of meta-analyses that is not representative of the field at large. Our non-systematic literature search and study selection strategy can only have exasperated this issue. It is also possible that our eligibility criteria correlated with the quality of the meta-analyses we reviewed. Second, although we had a mechanism in place to minimise the probability of data extraction errors on our part when evaluating reproducibility, it cannot be excluded that potential errors when extracting data for other variables (e.g., for PRISMA adherence) influenced our results as data extraction and coding was not checked by others (Buscemi et al., 2006; Jones et al., 2005).

Third, we focused on reproducibility of data extraction and calculation. Our study did not investigate reproducibility of the search for primary studies. This, is however, also an important aspect of reproducibility. Fourth, throughout the process of ES reproduction, we had to make subjective decisions that cannot be guaranteed to have been faultless. Notably, it was not always trivial to judge whether our procedure for reproducing a certain primary ES strictly followed the procedure purported to have been used by the meta-analysts. For example, we managed to approximate ES no. 5 in meta-analysis 3 by averaging the means and SDs of two outcomes and computing an ES based on the average value. We subsequently classified this ES as faithfully reproducible because the meta-analysts reported having used what each primary study defined as its primary outcome and this specific primary study defined both these outcomes as its primary outcomes. However, since the meta-analysts did not provide any information on how they computed the ES or any indication that they took an average, it is almost certain that they calculated the ES differently because otherwise we would have successfully reproduced the ES to the third decimal like we did the other brute-force reproducible ones. Other such examples are documented in the reproducibility report available on the project’s GitHub repo (taymalsalti.github.io/tDCS_meta-analysis/02_reproducibility_report.html).

Future similar works may hence aim for a more fine-grained and nuanced evaluation of reporting quality which goes beyond the minimal requirements set by reporting guidelines, a more comprehensive reproducibility testing which is not restricted to data extraction and ES calculation as well as a thorough investigation of robustness towards changes in analytical decisions (especially data selection and outcomes used), and a more principled approach towards evaluating publication bias assessment.

### Implications

All in all, our results indicate that the methodological limitations prevalent in meta-analyses in neighbouring fields can also be observed in meta-analyses studying the effect of tDCS on motor learning. Given the high status that meta-analyses enjoy on the “hierarchy of evidence” (Evans, 2003), they are likely to be influential both within and beyond the restricted realm of scientific publishing. For example, naive consumers of these meta-analyses might be misled into believing that they provide a definitive “proof” of tDCS’ effectiveness. Clinicians could rely on them in informing their decisions about treating patients.

The three meta-analyses we reviewed have been cited over 300 times^8^, which indicates they might be already heavily influencing researchers and other parties interested in tDCS effects. We argue that results of such meta-analyses need to be viewed with caution and in light of their limitations. Clinical guidelines citing evidence reported by such meta-analyses should appropriately caveat such citations for practitioners with detailed and accessible discussions about the uncertainties arising from their limitations. Transparent reporting and discussion of limitations on part of meta-analysts would facilitate the task of evaluating the strength of the evidence they deliver.

### Conclusions

We hope to have provided consumers of tDCS-motor learning research with an incentive to evaluate meta-analyses in this field more critically when making decisions about the use of tDCS and meta-analysts with aspects to consider when conducting meta-analyses in the future. By no means do we want to imply that the solutions presented above are easy. Despite the abundance and comprehensiveness of guidelines and tutorials on how to conduct a transparent and reproducible meta-analysis (e.g., Moreau & Gamble, 2020; Quintana, 2015), it is undeniable that meticulous adherence to guidelines and making one’s meta-analysis reproducible require a substantial amount of time and effort. Likewise, it cannot be expected from substantive researchers to be adept at every methodological/statistical aspect related to conducting a meta-analysis.

Guidelines, which are constantly being updated and which also include advancements in statistical procedures for controlling publication bias, may aid researchers in conducting meta-analyses (e.g., Vevea et al., 2019). Placing more emphasis on flagship open science practices such as pre-registration and data and code sharing (Maassen et al., 2020; Page, Moher, et al., 2021), may also help advance meta-analysis. Providing data and code facilitates reanalysis of the data and allows for analyses that investigate the impact of different methodological decisions and sensitivity of the results to them (Taylor & Munafò, 2016; e.g., Voracek et al., 2019). Potential measures to improve the situation on a higher, more structural level are ways to incentivise transparency-related practices (Bakker et al., 2012; Higginson & Munafò, 2016; Nosek et al., 2015) or journals employing specialised statistical review (Hardwicke et al., 2019).

Finally, since meta-analyses can only be as good as the primary literatures they synthesise, meta-analysts cannot be expected to carry all the responsibility for improvement (Aguinis, Pierce, et al., 2011; Borenstein et al., 2009). In the quest for a reliable cumulative science, efforts to counteract or alleviate the methodological issues found in the tDCS primary literature (e.g., neglected heterogeneity in employed tDCS parameters, compromised reproducibility due to incomplete reporting; Buch et al., 2017) must also be supported. Such efforts include checklists outlining all aspects which should be disclosed when reporting the results of a tDCS study (Buch et al., 2017) and statistical models which can be used to systematically determine the optimal tDCS parameters to use depending on the specific setting (Lipka et al., 2021). A more coherent primary body of literature is bound to lead to more informative synthesis.

## Data Availability

All data and code necessary for reproducing our results are available on https://github.com/TaymAlsalti/tDCS_meta-analysis

## Acknowledgement

We thank Esther Maassen for her generous support and input at various stages of this project.

## Supplemental Materials to the manuscript

**Table S1.** Reproducibility of primary SMDs, Meta-analysis 1.

| SMD no. | Reported SMD | Faithfully Reproduced SMD | Reproducibility classification | Reason for irreproducibility or approximation |
| --- | --- | --- | --- | --- |
| 1 | 0.16 | 0.16 | Faithfully reproducible | Not applicable. |
| 2 | 0.18 | 0.17 | Faithfully approximated | Values extracted from figure. Crossover design. Used total sample size for both the control and treatment group sample sizes. |
| 3 | 0.36 | 0.36 | Faithfully reproducible | Not applicable. |
| 4 | 0.08 | 0.12 | Faithfully approximated | Values extracted from figure. |
| 5 | 0.38 | 0.38 | Faithfully reproducible | Not applicable. |
| 6 | 0.04 | 0.04 | Faithfully reproducible | Not applicable. |
| 7 | 0.06 | 0.06 | Faithfully reproducible | Not applicable. |
| 8 | 1.59 | 1.68 | Faithfully irreproducible | Not inferable. |
| 9 | 1.08 | 1.15 | Faithfully irreproducible | Not inferable. |
| 10 | 1.05 | 1.02 | Faithfully approximated | Values extracted from figure. |

M-A METHODS IN TDCS - MOTOR LEARNING RESEARCH
| SMD no. | Reported SMD | Faithfully Reproduced SMD | Reproducibility classification | Reason for irreproducibility or approximation |
| --- | --- | --- | --- | --- |
| 11 | 1.39 | 1.35 | Faithfully approximated | Values extracted from figure. |
| 12 | 0.93 | NA | Brute-force irreproducible | Not inferable. Crossover design. |
| 13 | 0.82 | NA | Brute-force irreproducible | Not inferable. Crossover design. |
| 14 | 0.29 | 0.29 | Faithfully reproducible | Not applicable. |
| 15 | 0.61 | 0.64 | Brute-force approximated | Approximated using percentage change from baseline values. |
| 16 | 1.43 | 0.18 | Faithfully irreproducible, Brute-force reproducible | Outcome used does not correspond to description. |
| 17 | 0.94 | 0.18 | Faithfully irreproducible | Not inferable. |
| 18 | 0.24 | 0.54 | Faithfully irreproducible | Not inferable. |
| 19 | 0.65 | NA | Brute-force reproducible | Outcome used does not correspond to description. |
| 20 | 0.72 | NA | Brute-force reproducible | Successfully reproduced using a p-value derived from a medians test. |
| 21 | 0.53 | 1.61 | Faithfully irreproducible, brute-force irreproducible | Not inferable. |

**Table S2.**
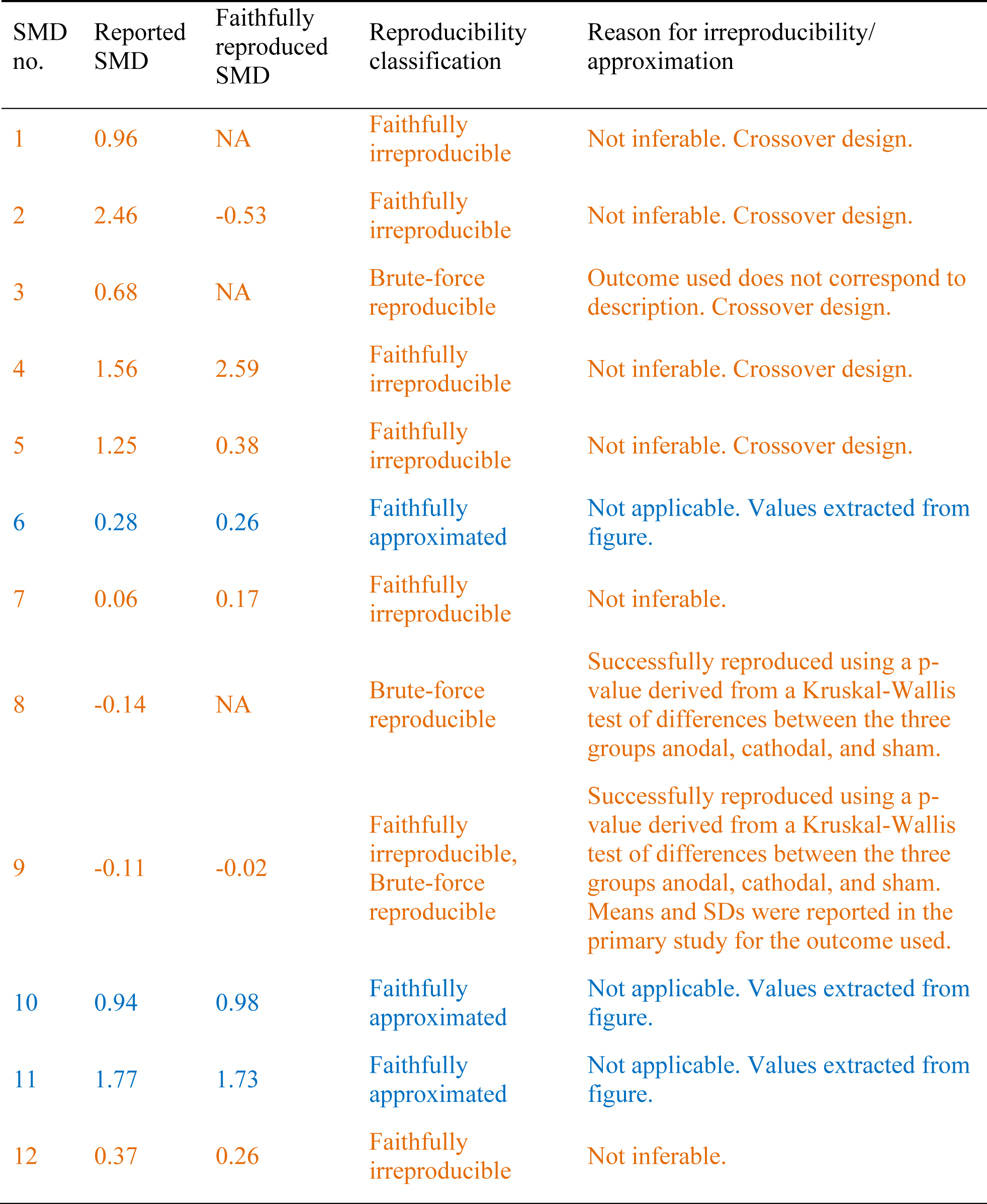
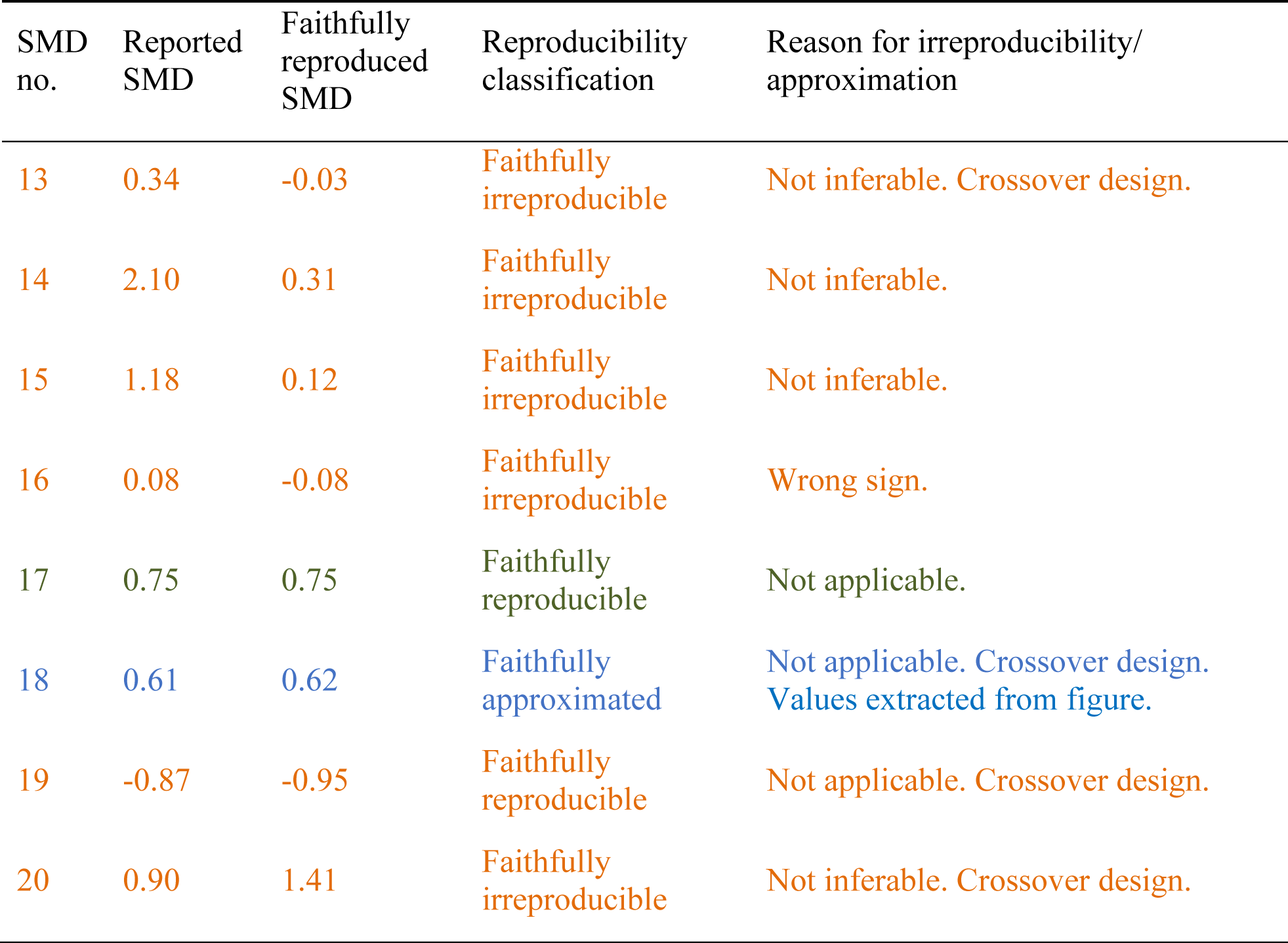
Reproducibility of primary SMDs, Meta-analysis 2.

**Table S3.** Reproducibility of primary SMDs, Meta-analysis 3.

| SMD no. | Reported SMD | Reproduced SMD | Reproducibility classification | Reason for irreproducibility |
| --- | --- | --- | --- | --- |
| 1 | 0.84 | NA | Brute-force reproducible | Successfully reproduced using a p-value (reported in the primary study as a range “<0.01”) derived from a difference in medians test. |
| 2 | 0.25 | 0.27 | Faithfully irreproducible, Brute-force approximated | Successfully approximated using the values for one of the two outcomes indicated to have been used and doubling the tDCS group sample size. |
| 3 | 0.98 | NA | Brute-force reproducible | Successfully reproduced using a p-value (reported as a range “<0.01”) derived from a medians test in combination with the total sample size in place of both treatment and control group |

M-A METHODS IN TDCS - MOTOR LEARNING RESEARCH
| SMD no. | Reported SMD | Reproduced SMD | Reproducibility classification | Reason for irreproducibility |
| --- | --- | --- | --- | --- |
|  |  |  |  | sample sizes. Notably, this was not a journal article, but a conference abstract. |
| 4 | 1.19 | NA | Brute-force reproducible | Outcome used does not correspond to description. |
| 5 | 0.59 | 0.56 | Faithfully approximated | Approximated by averaging two sets of means and SDs for two different outcomes. Although the meta-analysts did not describe having done this, we tenuously classified this reproduction as faithful because both outcomes were defined as primary outcomes in the primary study and the meta-analysts wrote that they used whatever the primary studies defined as primary outcomes. |
| 6 | 0.58 | NA | Brute-force reproducible | Successfully reproduced using a p-value based on difference between tDCS and sham groups in change from baseline |

### Supplementary Note 1

Besides coding whether any statistical methods were used at all and which, we tested for publication bias in each meta-analysis using 3 different methods: PET-PEESE (T. Stanley, 2008; T. D. Stanley & Doucouliagos, 2014), *p*-curve (Simonsohn et al., 2014b, 2014a), and the three-parameter selection model (McShane et al., 2016). For these analyses, we used the ESs and the sample sizes reported in the meta-analyses along with the sampling variances extracted from the funnel plots in the case of the first two meta-analyses and calculated based on the CIs in the case of the third meta-analysis. We chose these three methods specifically for practical reasons: they are implemented in already available R packages (see https://taymalsalti.github.io/tDCS_meta-analysis/03_pub-bias_outlier_analyses.html for more details).

For meta-analysis 1, the estimate of the true effect produced by the three-parameter selection model (0.59) was virtually identical to the original. The estimate produced by the *p*-curve was larger (0.70). Only the PET-PEESE intercepts (0.34 and 0.45, respectively) indicated that the random-effects model-based estimates might be overestimating the true effect. For meta-analysis 2, the selection model (0.20), *p*-curve (0.18), and PEESE (0.21) estimates were much smaller than the original (0.62). The PET intercept (−0.12) was negative. Similar results were observed for the last meta-analysis: the estimates produced by the the *p*-curve and PET-PEESE were 0.40, −0.71, and 0.01, respectively. Only the selection model yielded an estimate which is close to the one based on the random-effects model (0.62). These results indicate that the meta-analysts’ conclusion that publication bias is not a concern are not robust, but rather sensitive to the specific method used.

**Figure S1.**
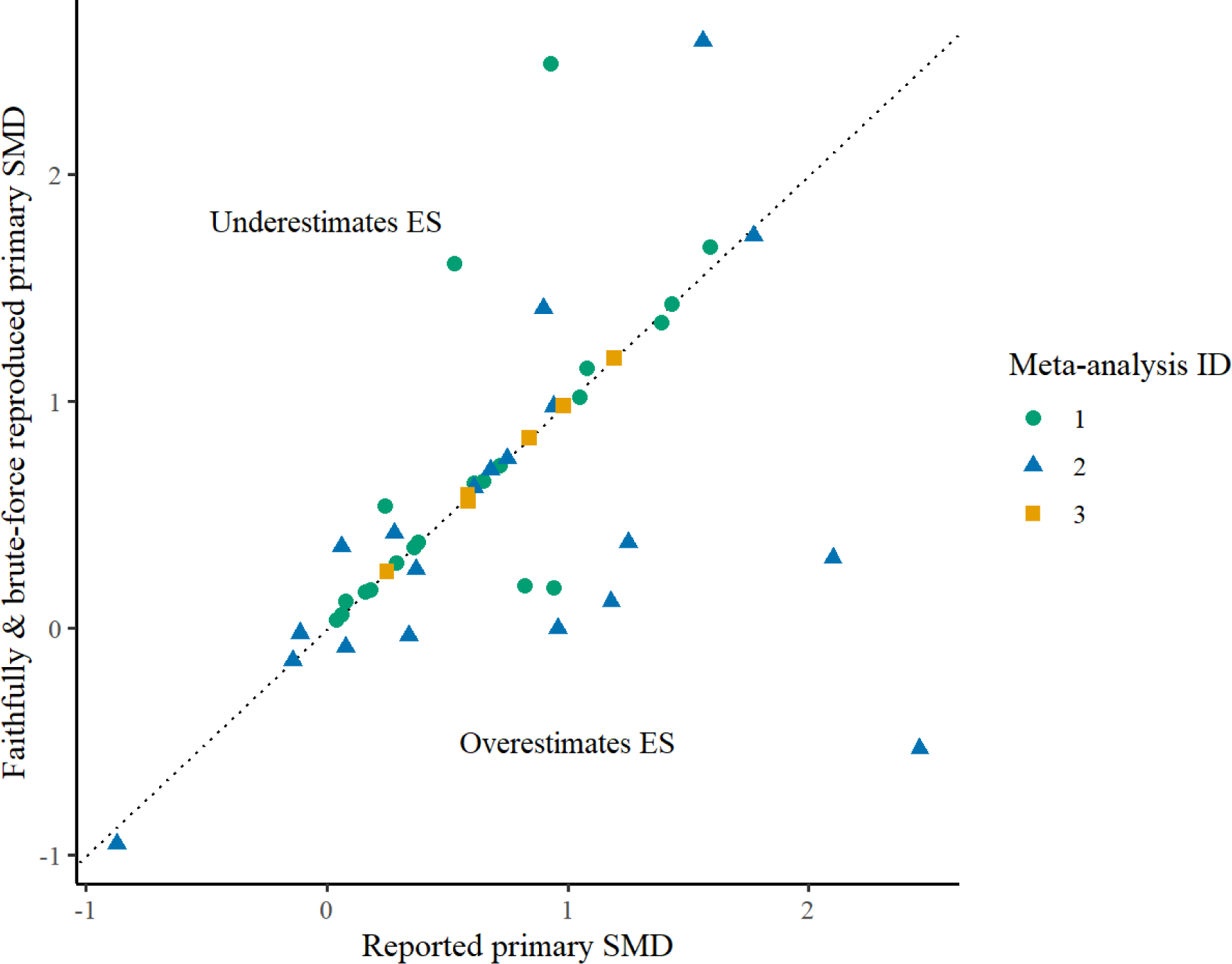
Including brute-force reproduced SMDs

### Outlier analyses

**Figure S2.**
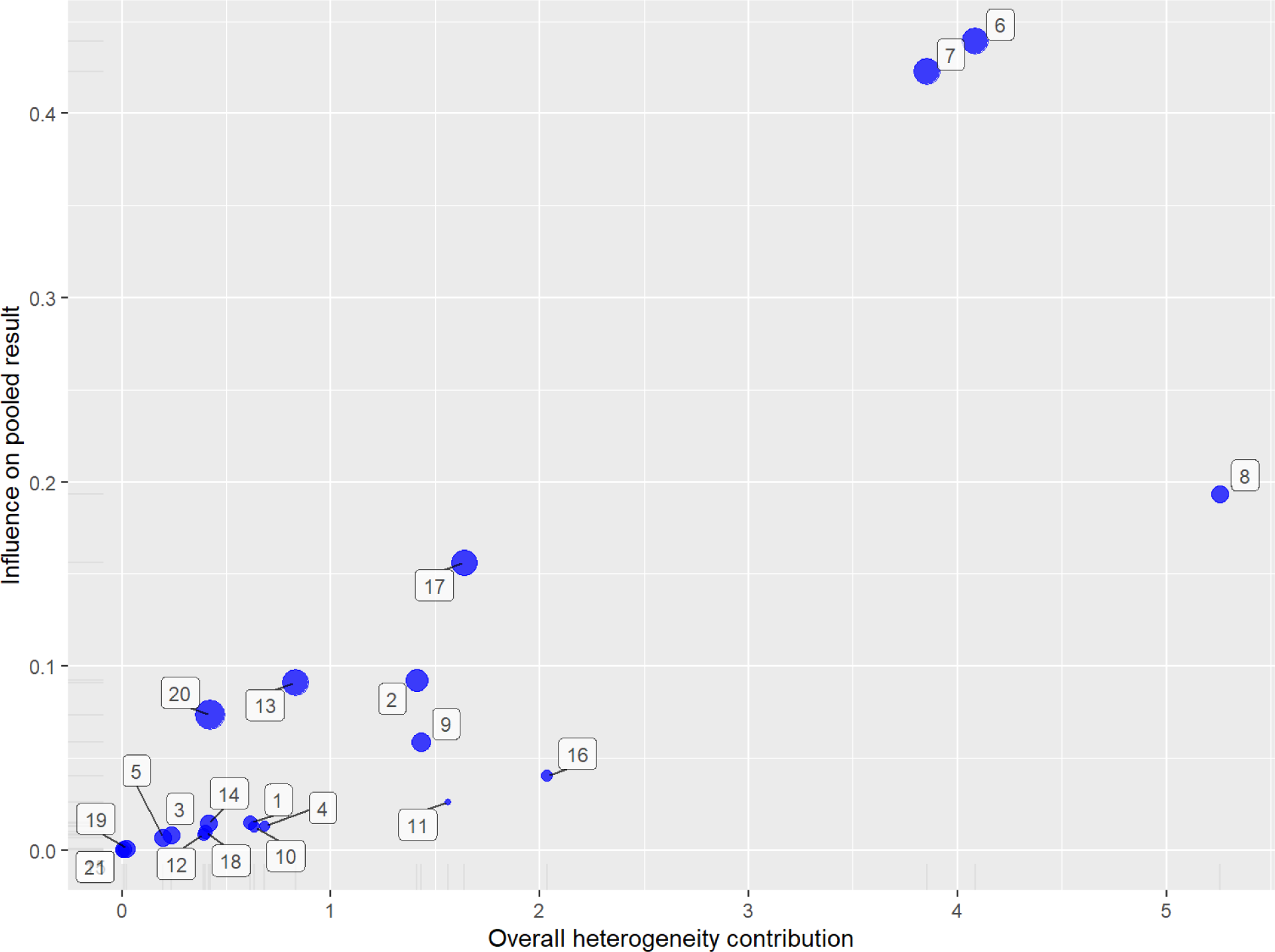
Outlier analysis, meta-analysis 1. ESs 6, 7, and 8 contribute disproportionately to both the variance and the pooled ES.

**Figure S3.**
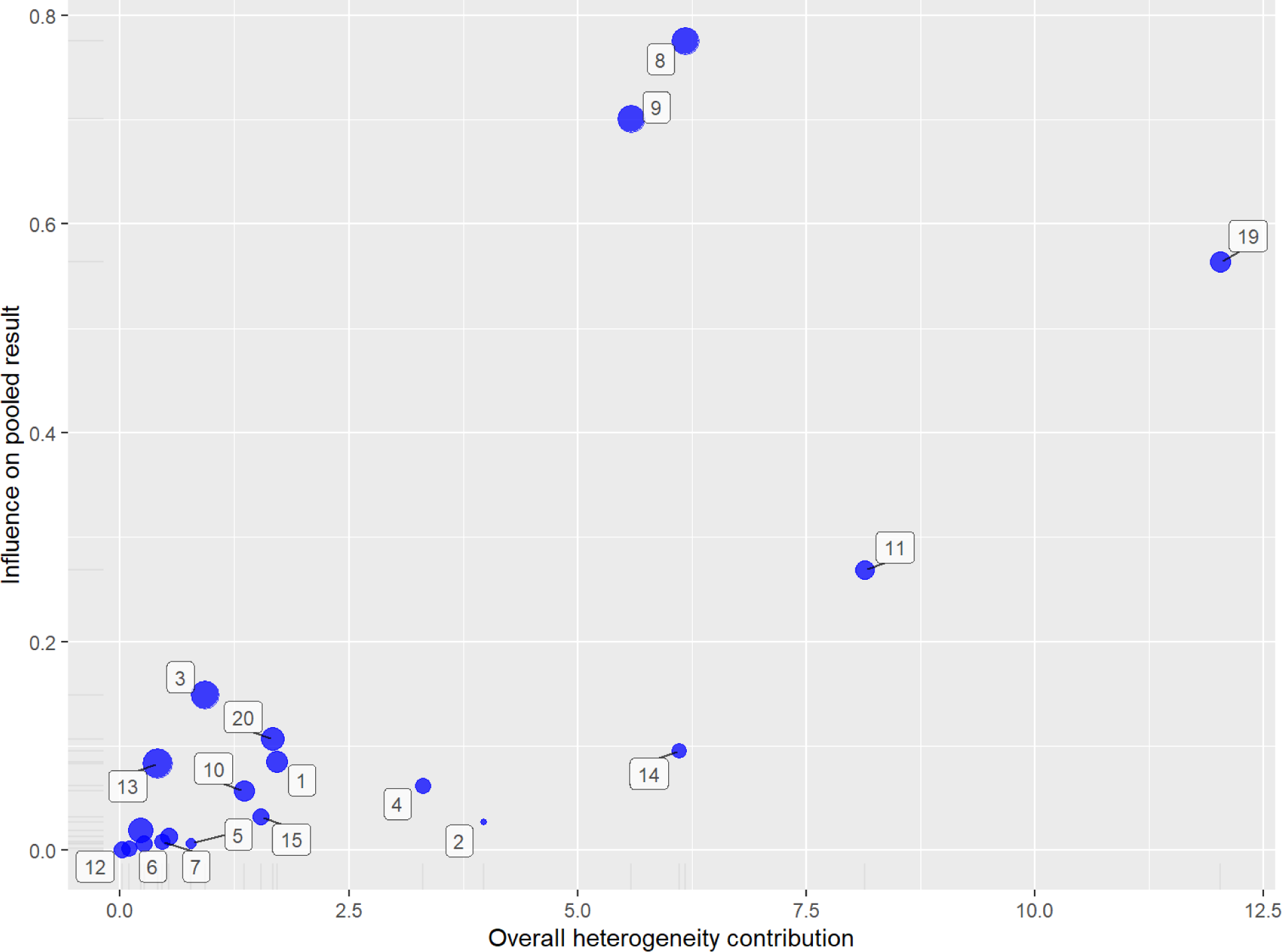
Outlier analysis, meta-analysis 2. ES 19 contributes disproportionately to both the variance and the pooled ES. ESs 8 and 9 mostly to the pooled ES.

**Figure S4.**
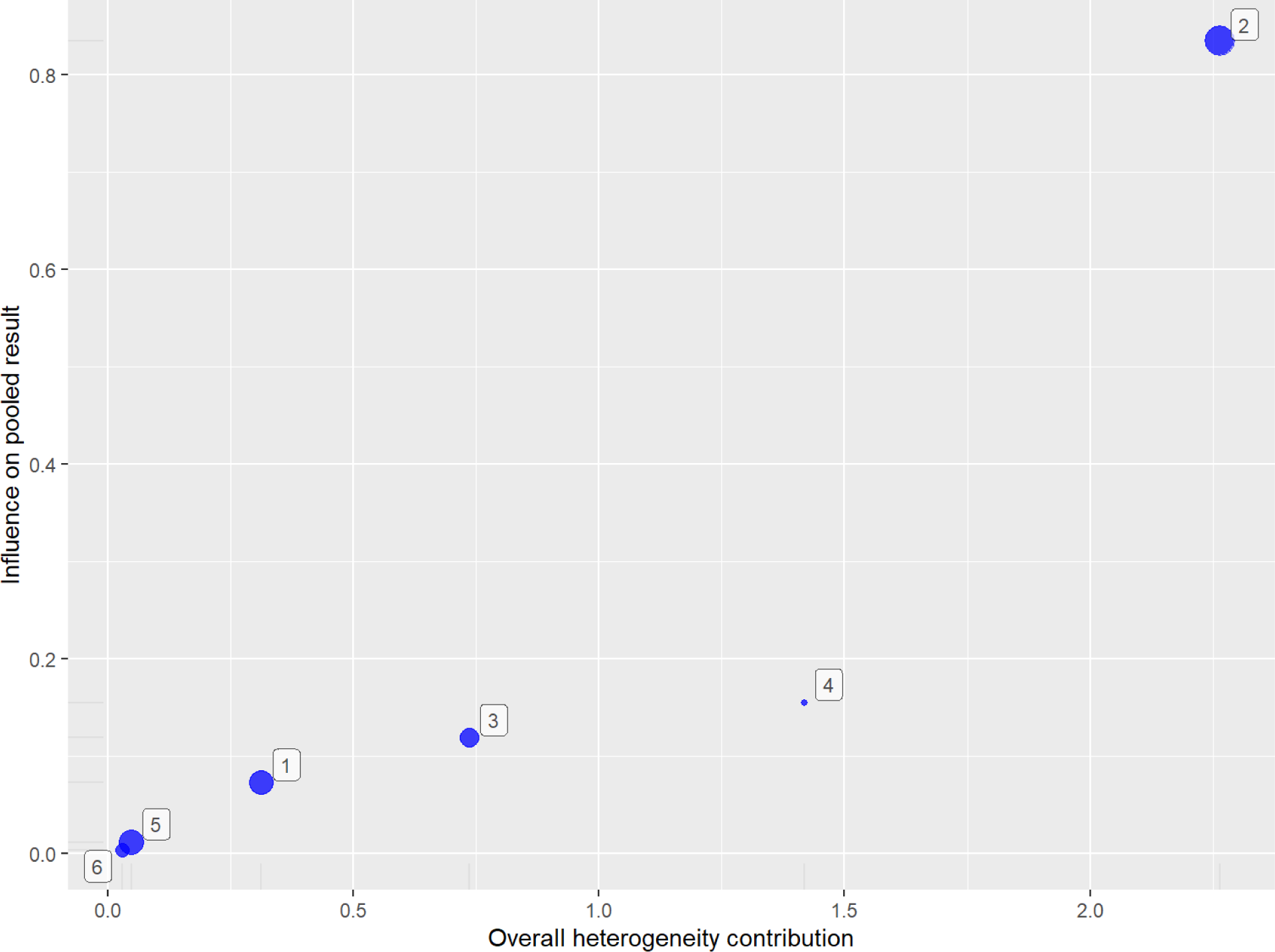
Outlier analysis, meta-analysis 3. ES 19 contributes disproportionately to both the variance and the pooled ES.

1 One review of meta-analyses in the organisational sciences found that 21 different analytical decisions had a negligible impact on the pooled effect size estimate (Aguinis, Dalton, et al., 2011)

2 E.g., for 4 out of the 8 meta-analyses Ford et al. (2010) reviewed, the reproduced pooled ES estimates were no longer significant. Similarly, one of the meta-analyses Gøtzsche et al. (2007) reviewed was later retracted and two were no longer significant in their reproduced versions.

3 Along with the corresponding sampling variances/SEs. Since none of the meta-analyses reported how they calculated these values, we simply calculated the sampling variances that correspond to the type of ES we calculated for any given primary study. In general, the sampling variances were numerically reproducible whenever their corresponding primary ES was reproducible.

4 This is because we had to extract some values (e.g., means and CIs) from figures reported in the primary studies, which is bound to lead to rounding deviations if the meta-analysts did so, too.

5 Sampling variances were not reported in any of the three meta-analyses. They were calculated based on SEs extracted from funnel plots in the case of the first two meta-analyses and based on CIs for the third meta-analysis.

6 In the case of the first meta-analysis, we were informed by the first author of the meta-analysis that they used the Comprehensive Meta-Analysis software in response to an email asking for further information. Our email’s text as well as all the reproducibility-relevant information contained in the meta-analysts’ response (not the actual text of their responses) can be found in the document “Email_to_authors” on our OSF project.

7 Namely that they used the outcome the primary study defined as their primary outcome.

8 296, 24, and 14 times, respectively, as of 05.06.2024, according to Google Scholar. The first meta-analysis was cited by Lefaucheur et al. (2017), one of the papers outlining guidelines for the clinical use of tDCS we cited above.

## References

1. Ada, S., Sharman, R., & Balkundi, P. (2012). Impact of meta-analytic decisions on the conclusions drawn on the business value of information technology. Decision Support Systems, 54(1), 521–533. 10.1016/j.dss.2012.07.001

2. Aguinis, H., Dalton, D. R., Bosco, F. A., Pierce, C. A., & Dalton, C. M. (2011). Meta-Analytic Choices and Judgment Calls: Implications for Theory Building and Testing, Obtained Effect Sizes, and Scholarly Impact. Journal of Management, 37(1), 5–38. 10.1177/0149206310377113

3. Aguinis, H., Pierce, C. A., Bosco, F. A., Dalton, D. R., & Dalton, C. M. (2011). Debunking Myths and Urban Legends About Meta-Analysis. Organizational Research Methods, 14(2), 306–331. 10.1177/1094428110375720

4. Austin, A., Jiga-Boy, G. M., Rea, S., Newstead, S. A., Roderick, S., Davis, N. J., Clement, R. M., & Boy, F. (2016). Prefrontal Electrical Stimulation in Non-depressed Reduces Levels of Reported Negative Affects from Daily Stressors. Frontiers in Psychology, 7, 315. 10/gnvzvg

5. Bakker, M., van Dijk, A., & Wicherts, J. M. (2012). The Rules of the Game Called Psychological Science. Perspectives on Psychological Science, 7(6), 543–554. 10/f4fbz2

6. Balduzzi, S., Rücker, G., & Schwarzer, G. (2019). How to perform a meta-analysis with R: a practical tutorial. Evidence-Based Mental Health, 22, 153–160. 10/ggchpv

7. Bates, D., & Maechler, M. (2021). *Matrix: Sparse and dense matrix classes and methods* [Manual]. https://CRAN.R-project.org/package=Matrix

8. Bennabi, D., Pedron, S., Haffen, E., Monnin, J., Peterschmitt, Y., & Van Waes, V. (2014). Transcranial direct current stimulation for memory enhancement: From clinical research to animal models. Frontiers in Systems Neuroscience, 8, 159. 10/gdh996

9. Borenstein, M., Hedges, L., Higgins, J. P. T., & Rothstein, H. R. (2009). Criticisms of meta-analysis. In Introduction to Meta-Analysis (pp. 377–387). 10.1002/9780470743386.ch43

10. Borenstein, M., & Hedges, L. V. (2019). Effect sizes for meta-analyses. In The Handbook of Research Synthesis and Meta-Analysis, (pp. 207–241).

11. Broman, K., Cetinkaya-Rundel, M., Nussbaum, A., Paciorek, C., Peng, R., Turek, D., & Wickham, H. (2017). *Recommendations to Funding Agencies for Supporting Reproducible Research*. https://www.amstat.org/asa/files/pdfs/POL-ReproducibleResearchRecommendations.pdf

12. Brunoni, A. R., Moffa, A. H., Fregni, F., Palm, U., Padberg, F., Blumberger, D. M., Daskalakis, Z. J., Bennabi, D., Haffen, E., Alonzo, A., & Loo, C. K. (2016). Transcranial direct current stimulation for acute major depressive episodes: Meta-analysis of individual patient data. The British Journal of Psychiatry, 208(6), 522–531. 10/f8qtrg

13. Buch, E. R., Santarnecchi, E., Antal, A., Born, J., Celnik, P. A., Classen, J., Gerloff, C., Hallett, M., Hummel, F. C., Nitsche, M. A., Pascual-Leone, A., Paulus, W. J., Reis, J., Robertson, E. M., Rothwell, J. C., Sandrini, M., Schambra, H. M., Wassermann, E. M., Ziemann, U., & Cohen, L. G. (2017). Effects of tDCS on motor learning and memory formation: A consensus and critical position paper. Clinical Neurophysiology, 128(4), 589–603. 10.1016/j.clinph.2017.01.004

14. Buscemi, N., Hartling, L., Vandermeer, B., Tjosvold, L., & Klassen, T. P. (2006). Single data extraction generated more errors than double data extraction in systematic reviews. Journal of Clinical Epidemiology, 59(7), 697–703. 10.1016/j.jclinepi.2005.11.010

15. Carter, E. C., Schönbrodt, F. D., Gervais, W. M., & Hilgard, J. (2019). Correcting for Bias in Psychology: A Comparison of Meta-Analytic Methods. Advances in Methods and Practices in Psychological Science, 2(2), 115–144. 10.1177/2515245919847196

16. Charvet, L. E., Shaw, M. T., Bikson, M., Woods, A. J., & Knotkova, H. (2020). Supervised transcranial direct current stimulation (tDCS) at home: A guide for clinical research and practice. *Brain Stimulation: Basic*, Translational, and Clinical Research in Neuromodulation, 13(3), 686–693. 10.1016/j.brs.2020.02.011

17. Davis, N. J. (2016). The regulation of consumer tDCS: Engaging a community of creative self-experimenters. JL & Biosciences, 3, 304.

18. DerSimonian, R., & Laird, N. (1986). Meta-analysis in clinical trials. Controlled Clinical Trials, 7(3), 177–188. 10/fdpm2j

19. Evans, D. (2003). Hierarchy of evidence: A framework for ranking evidence evaluating healthcare interventions. Journal of Clinical Nursing, 12(1), 77–84. 10/dcv5w9

20. Ford, A. C., Guyatt, G. H., Talley, N. J., & Moayyedi, P. (2010). Errors in the conduct of systematic reviews of pharmacological interventions for irritable bowel syndrome. The American Journal of Gastroenterology, 105(2), 280–288. 10.1038/ajg.2009.658

21. Fregni, F., Nitsche, M. A., Loo, C. K., Brunoni, A. R., Marangolo, P., Leite, J., Carvalho, S., Bolognini, N., Caumo, W., Paik, N. J., Simis, M., Ueda, K., Ekhitari, H., Luu, P., Tucker, D. M., Tyler, W. J., Brunelin, J., Datta, A., Juan, C. H.,… Bikson, M. (2015). Regulatory Considerations for the Clinical and Research Use of Transcranial Direct Current Stimulation (tDCS): Review and recommendations from an expert panel. Clinical Research and Regulatory Affairs, 32(1), 22–35. 10/gnvzvf

22. Gazzaniga, M., Ivry, R. B., & Mangun, G. R. (2018). Methods of Cognitive Neuroscience. In Cognitive Neuroscience: The Biology of the Mind (Fifth edition). W. W. Norton & Company.

23. Gebodh, N., Esmaeilpour, Z., Adair, D., Schestattsky, P., Fregni, F., & Bikson, M. (2019). Transcranial Direct Current Stimulation Among Technologies for Low-Intensity Transcranial Electrical Stimulation: Classification, History, and Terminology. In H. Knotkova, M. A. Nitsche, M. Bikson, & A. J. Woods (Eds.), Practical Guide to Transcranial Direct Current Stimulation: Principles, Procedures and Applications (pp. 3–43). Springer International Publishing. 10.1007/978-3-319-95948-1_1

24. Gianni, E., Bertoli, M., Simonelli, I., Paulon, L., Tecchio, F., & Pasqualetti, P. (2021). tDCS randomized controlled trials in no-structural diseases: A quantitative review. Scientific Reports, 11(1), 16311. 10/gnvzvm

25. Gleser, L. J., & Olkin, I. (2009). Stochastically dependent effect sizes. In The handbook of research synthesis and meta-analysis, 2nd ed (pp. 357–376). Russell Sage Foundation.

26. Goodman, S. N., Fanelli, D., & Ioannidis, J. P. A. (2016). What does research reproducibility mean? Science Translational Medicine, 8(341), 341ps12. 10/gc5sjs

27. Gopalakrishnan, S., & Ganeshkumar, P. (2013). Systematic Reviews and Meta-analysis: Understanding the Best Evidence in Primary Healthcare. Journal of Family Medicine and Primary Care, 2(1), 9–14. 10.4103/2249-4863.109934

28. Gøtzsche, P. C., Hróbjartsson, A., Marić, K., & Tendal, B. (2007). Data Extraction Errors in Meta-analyses That Use Standardized Mean Differences. JAMA, 298(4), 430–437. 10.1001/jama.298.4.430

29. Guzzo, R., Jackson, S., & Katzell, R. (1987). Meta-analysis analysis. Research in Organizational Behavior, 9, 407–442.

30. Hardwicke, T. E., Frank, M. C., Vazire, S., & Goodman, S. N. (2019). Should Psychology Journals Adopt Specialized Statistical Review? Advances in Methods and Practices in Psychological Science, 2(3), 240–249. 10/gf4mm5

31. Harrer, M., Cuijpers, P., Furukawa, T. A., & Ebert, D. D. (2021). Doing Meta-Analysis With R: A Hands-On Guide (1st ed.). Chapman & Hall/CRC Press. https://www.routledge.com/Doing-Meta-Analysis-with-R-A-Hands-On-Guide/Harrer-Cuijpers-Furukawa-Ebert/p/book/9780367610074

32. Harrer, M., Cuijpers, P., Furukawa, T., & Ebert, D. D. (2019). dmetar: Companion r package for the guide ‘doing meta-analysis in r’ [Manual]. http://dmetar.protectlab.org/

33. Henry, L., & Wickham, H. (2020). *purrr: Functional programming tools* [Manual]. https://CRAN.R-project.org/package=purrr

34. Higgins, J. P., Thomas, J., Chandler, J., Cumpston, M., Li, T., Page, M. J., & Welch, V. A. (2019). Cochrane handbook for systematic reviews of interventions. John Wiley & Sons.

35. Higginson, A. D., & Munafò, M. R. (2016). Current Incentives for Scientists Lead to Underpowered Studies with Erroneous Conclusions. PLOS Biology, 14(11), e2000995. 10/f9mzp4

36. Horvath, J. C., Forte, J. D., & Carter, O. (2015). Quantitative Review Finds No Evidence of Cognitive Effects in Healthy Populations From Single-session Transcranial Direct Current Stimulation (tDCS). Brain Stimulation, 8(3), 535–550. 10/zm9

37. Hoyt, A. C. D. R. & W. T. (2014). MAd: Meta-analysis with mean differences [Manual]. https://cran.r-project.org/package=MAd

38. Hozo, S. P., Djulbegovic, B., & Hozo, I. (2005). Estimating the mean and variance from the median, range, and the size of a sample. BMC Medical Research Methodology, 5(1), 13. 10/dt5pn6

39. Hung, C.-M., Zeng, B.-Y., Zeng, B.-S., Sun, C.-K., Cheng, Y.-S., Su, K.-P., Wu, Y.-C., Chen, T.-Y., Lin, P.-Y., Liang, C.-S., Hsu, C.-W., Chu, C.-S., Chen, Y.-W., Wu, M.-K., & Tseng, P.-T. (2021). The Efficacy of Transcranial Direct Current Stimulation in Enhancing Surgical Skill Acquisition: A Preliminary Meta-Analysis of Randomized Controlled Trials. Brain Sciences, 11(6), Article 6. 10.3390/brainsci11060707

40. Inzlicht, M., Gervais, W., & Berkman, E. (2015). Bias-Correction Techniques Alone Cannot Determine Whether Ego Depletion is Different from Zero: Commentary on Carter, Kofler, Forster, & McCullough, 2015. 10.2139/ssrn.2659409

41. Ioannidis, J. P. A. (2016). The Mass Production of Redundant, Misleading, and Conflicted Systematic Reviews and Meta-analyses. The Milbank Quarterly, 94(3), 485–514. 10.1111/1468-0009.12210

42. Jones, A. P., Remmington, T., Williamson, P. R., Ashby, D., & Smyth, R. L. (2005). High prevalence but low impact of data extraction and reporting errors were found in Cochrane systematic reviews. Journal of Clinical Epidemiology, 58(7), 741–742. 10.1016/j.jclinepi.2004.11.024

43. Kang, N., Summers, J. J., & Cauraugh, J. H. (2015). Transcranial direct current stimulation facilitates motor learning post-stroke: A systematic review and meta-analysis. *Journal of Neurology*, Neurosurgery & Psychiatry. https://jnnp.bmj.com/content/jnnp/early/2015/08/28/jnnp-2015-311242.full.pdf?casa_token=2G7nyIjiF-EAAAAA:2wvvN4-GWGW2u_8xp_dKvVduXwX_wvDoW5QaL3nAo_IQp-DzhNQS1jxvB6IJJtdCJGjx3OdAwVs

44. Kang, N., Summers, J. J., & Cauraugh, J. H. (2016). Transcranial direct current stimulation facilitates motor learning post-stroke: A systematic review and meta-analysis. In JOURNAL OF NEUROLOGY NEUROSURGERY AND PSYCHIATRY (Vol. 87, Issue 4, pp. 345–355). BMJ PUBLISHING GROUP. 10.1136/jnnp-2015-311242

45. Kang, N., Weingart, A., & Cauraugh, J. H. (2018). Transcranial direct current stimulation and suppression of contralesional primary motor cortex post-stroke: A systematic review and meta-analysis. Brain Injury, 32(9), 1063–1070. 10/gf94wc

46. Lakens, D., Hilgard, J., & Staaks, J. (2016). On the reproducibility of meta-analyses: Six practical recommendations. BMC Psychology, 4(1), 24. 10.1186/s40359-016-0126-3

47. Lakens, D., Page-Gould, E., Assen, M. A. L. M. van, Spellman, B., Schönbrodt, F., Hasselman, F., Corker, K. S., Grange, J. A., Sharples, A., Cavender, C., Augusteijn, H., Augusteijn, H., Gerger, H., Locher, C., Miller, I. D., Anvari, F., & Scheel, A. M. (2017). Examining the Reproducibility of Meta-Analyses in Psychology: A Preliminary Report. 10.31222/osf.io/xfbjf

48. Lefaucheur, J.-P. (2016). A comprehensive database of published tDCS clinical trials (2005-2016). In NEUROPHYSIOLOGIE CLINIQUE-CLINICAL NEUROPHYSIOLOGY (Vol. 46, Issue 6, pp. 319–398). ELSEVIER FRANCE-EDITIONS SCIENTIFIQUES MEDICALES ELSEVIER. 10.1016/j.neucli.2016.10.002

49. Lefaucheur, J.-P., Antal, A., Ayache, S. S., Benninger, D. H., Brunelin, J., Cogiamanian, F., Cotelli, M., De Ridder, D., Ferrucci, R., Langguth, B., Marangolo, P., Mylius, V., Nitsche, M. A., Padberg, F., Palm, U., Poulet, E., Priori, A., Rossi, S., Schecklmann, M.,… Paulus, W. (2017). Evidence-based guidelines on the therapeutic use of transcranial direct current stimulation (tDCS). Clinical Neurophysiology, 128(1), 56–92. 10.1016/j.clinph.2016.10.087

50. Liberati, A., Altman, D. G., Tetzlaff, J., Mulrow, C., Gøtzsche, P. C., Ioannidis, J. P. A., Clarke, M., Devereaux, P. J., Kleijnen, J., & Moher, D. (2009). The PRISMA statement for reporting systematic reviews and meta-analyses of studies that evaluate healthcare interventions: Explanation and elaboration. BMJ, 339, b2700. 10.1136/bmj.b2700

51. Lipka, R., Ahlers, E., Reed, T. L., Karstens, M. I., Nguyen, V., Bajbouj, M., & Cohen Kadosh, R. (2021). Resolving heterogeneity in transcranial electrical stimulation efficacy for attention deficit hyperactivity disorder. Experimental Neurology, 337, 113586. 10/gnvzvd

52. Liu, Y., Gu, N., Cao, X., Zhu, Y., Wang, J., Smith, R. C., & Li, C. (2021). Effects of transcranial electrical stimulation on working memory in patients with schizophrenia: A systematic review and meta-analysis. Psychiatry Research, 296, 113656. 10/gnvzvh

53. Luedtke, K., Rushton, A., Wright, C., Geiss, B., Juergens, T. P., & May, A. (2012). Transcranial Direct Current Stimulation for the Reduction of Clinical and Experimentally Induced Pain: A Systematic Review and Meta-analysis. The Clinical Journal of Pain, 28(5), 452–461. 10/gnvzvk

54. Maassen, E., Assen, M. A. L. M. van, Nuijten, M. B., Olsson-Collentine, A., & Wicherts, J. M. (2020). Reproducibility of individual effect sizes in meta-analyses in psychology. PLOS ONE, 15(5), e0233107. 10.1371/journal.pone.0233107

55. Madeyski, L., & Kitchenham, B. (2018). Effect sizes and their variance for AB/BA crossover design studies. Empirical Software Engineering, 23(4), 1982–2017. 10/gdst65

56. McShane, B. B., Böckenholt, U., & Hansen, K. T. (2016). Adjusting for Publication Bias in Meta-Analysis: An Evaluation of Selection Methods and Some Cautionary Notes. Perspectives on Psychological Science, 11(5), 730–749. 10.1177/1745691616662243

57. Moher, Cook, D. J., Eastwood, S., Olkin, I., Rennie D. & Stroup, D. F. (2000). Improving the Quality of Reports of Meta-Analyses of Randomised Controlled Trials: The QUOROM Statement. Oncology Research and Treatment, 23(6), 597–602. 10/ckwqz5

58. Moher, Liberati, A., Tetzlaff, J., Altman D. G., & Group T.P. (2009). Preferred Reporting Items for Systematic Reviews and Meta-Analyses: The PRISMA Statement. PLOS Medicine, 6(7), e1000097. 10/bq3jpc

59. Moreau, D., & Gamble, B. (2020). Conducting a meta-analysis in the age of open science: Tools, tips, and practical recommendations. Psychological Methods, No Pagination Specified-No Pagination Specified. 10/gjfpv6

60. Morganti, A. (2007). The impact of meta-analyses on clinical practice: The benefits. Journal of Nephrology, 20 *Suppl 12*, S1–3.

61. Morris, S. B., & DeShon, R. P. (2002). Combining effect size estimates in meta-analysis with repeated measures and independent-groups designs. Psychological Methods, 7(1), 105–125. 10.1037/1082-989X.7.1.105

62. Nitsche, M. A., Cohen, L. G., Wassermann, E. M., Priori, A., Lang, N., Antal, A., Paulus, W., Hummel, F., Boggio, P. S., Fregni, F., & Pascual-Leone, A. (2008). Transcranial direct current stimulation: State of the art 2008. Brain Stimulation, 1(3), 206–223. 10/fd6s2h

63. Nosek, B. A., Alter, G., Banks, G. C., Borsboom, D., Bowman, S. D., Breckler, S. J., Buck, S., Chambers, C. D., Chin, G., Christensen, G., Contestabile, M., Dafoe, A., Eich, E., Freese, J., Glennerster, R., Goroff, D., Green, D. P., Hesse, B., Humphreys, M.,… Yarkoni, T. (2015). Promoting an open research culture. Science, 348(6242), 1422–1425. 10/gcpzwn

64. Page, M. J., McKenzie, J. E., Bossuyt, P. M., Boutron, I., Hoffmann, T. C., Mulrow, C. D., Shamseer, L., Tetzlaff, J. M., Akl, E. A., Brennan, S. E., Chou, R., Glanville, J., Grimshaw, J. M., Hróbjartsson, A., Lalu, M. M., Li, T., Loder, E. W., Mayo-Wilson, E., McDonald, S.,… Moher, D. (2021). The PRISMA 2020 statement: An updated guideline for reporting systematic reviews. BMJ, 372, n71. 10.1136/bmj.n71

65. Page, M. J., & Moher, D. (2017). Evaluations of the uptake and impact of the Preferred Reporting Items for Systematic reviews and Meta-Analyses (PRISMA) Statement and extensions: A scoping review. Systematic Reviews, 6(1), 263. 10.1186/s13643-017-0663-8

66. Page, M. J., Moher, D., Bossuyt, P. M., Boutron, I., Hoffmann, T. C., Mulrow, C. D., Shamseer, L., Tetzlaff, J. M., Akl, E. A., Brennan, S. E., Chou, R., Glanville, J., Grimshaw, J. M., Hróbjartsson, A., Lalu, M. M., Li, T., Loder, E. W., Mayo-Wilson, E., McDonald, S.,… McKenzie, J. E. (2021). PRISMA 2020 explanation and elaboration: Updated guidance and exemplars for reporting systematic reviews. BMJ, 372, n160. 10.1136/bmj.n160

67. Polanin, J. R., Hennessy, E. A., & Tsuji, S. (2020). Transparency and Reproducibility of Meta-Analyses in Psychology: A Meta-Review. Perspectives on Psychological Science, 15(4), 1026–1041. 10.1177/1745691620906416

68. Quintana, D. S. (2015). From pre-registration to publication: A non-technical primer for conducting a meta-analysis to synthesize correlational data. Frontiers in Psychology, 6, 1549. 10/f3jrqf

69. R Core Team. (2021). R: A language and environment for statistical computing [Manual]. https://www.R-project.org/

70. Renkewitz, F., & Keiner, M. (2019). How to Detect Publication Bias in Psychological Research. Zeitschrift Für Psychologie, 227(4), 261–279. 10.1027/2151-2604/a000386

71. Rohatgi, A. (2021). WebPlotDigitizer [Computer software]. https://wpd.starrydata2.org/

72. Rosen, D. S., Erickson, B., Kim, Y. E., Mirman, D., Hamilton, R. H., & Kounios, J. (2016). Anodal tDCS to Right Dorsolateral Prefrontal Cortex Facilitates Performance for Novice Jazz Improvisers but Hinders Experts. Frontiers in Human Neuroscience, 10, 579. 10/gf932m

73. Schalken, N., & Rietbergen, C. (2017). The Reporting Quality of Systematic Reviews and Meta-Analyses in Industrial and Organizational Psychology: A Systematic Review. Frontiers in Psychology, 8. 10.3389/fpsyg.2017.01395

74. Simmons, J. P., Nelson, L. D., & Simonsohn, U. (2011). False-Positive Psychology: Undisclosed Flexibility in Data Collection and Analysis Allows Presenting Anything as Significant. Psychological Science, 22(11), 1359–1366. 10/bxbw3c

75. Taylor, A. E., & Munafò, M. R. (2016). Triangulating meta-analyses: The example of the serotonin transporter gene, stressful life events and major depression. BMC Psychology, 4(1), 23. 10.1186/s40359-016-0129-0

76. Tobias, A. (1999). Assessing the influence of a single study in the meta-anyalysis estimate. Stata Technical Bulletin, 8(47). https://econpapers.repec.org/article/tsjstbull/y_3a1999_3av_3a8_3ai_3a47_3asbe26.htm

77. Valentine, J. C., Pigott, T. D., & Rothstein, H. R. (2010). How Many Studies Do You Need?: A Primer on Statistical Power for Meta-Analysis. Journal of Educational and Behavioral Statistics, 35(2), 215–247. 10.3102/1076998609346961

78. van Aert, R. C. M., Wicherts, J. M., & van Assen, M. A. L. M. (2016). Conducting Meta-Analyses Based on p Values: Reservations and Recommendations for Applying p-Uniform and p-Curve. Perspectives on Psychological Science, 11(5), 713–729. 10.1177/1745691616650874

79. Vevea, J. L., Coburn, K., & Sutton, A. (2019). Publication Bias. In H. Cooper, L. V. Hedges, & J. C. Valentine (Eds.), The Handbook of Research Synthesis and Meta-Analysis (pp. 383–430). Russell Sage Foundation. 10.7758/9781610448864.21

80. Viechtbauer, W. (2010). Conducting meta-analyses in R with the metafor package. Journal of Statistical Software, 36(3), 1–48. 10/gckfpj

81. Voracek, M., Kossmeier, M., & Tran, U. S. (2019). Which data to meta-analyze, and how? A specification-curve and multiverse-analysis approach to meta-analysis. Zeitschrift Für Psychologie, 227(1), 64–82. 10.1027/2151-2604/a000357

82. Wan, X., Wang, W., Liu, J., & Tong, T. (2014). Estimating the sample mean and standard deviation from the sample size, median, range and/or interquartile range. BMC Medical Research Methodology, 14(1), 135. 10/f7xmgr

83. Weissgerber, S. C., Brunmair, M., & Rummer, R. (2021). Null and Void? Errors in Meta-analysis on Perceptual Disfluency and Recommendations to Improve Meta-analytical Reproducibility. Educational Psychology Review, 33(3), 1221–1247. 10.1007/s10648-020-09579-1

84. Wexler, A. (2018). Who Uses Direct-to-Consumer Brain Stimulation Products, and Why? A Study of Home Users of tDCS Devices. Journal of Cognitive Enhancement, 2(1), 114–134. 10.1007/s41465-017-0062-z

85. Wickham, H. (2016). ggplot2: Elegant graphics for data analysis. Springer-Verlag New York. https://ggplot2.tidyverse.org

86. Wickham, H. (2021). *tidyr: Tidy messy data* [Manual]. https://CRAN.R-project.org/package=tidyr

87. Wickham, H., François, R., Henry, L., & Müller, K. (2021). dplyr: A grammar of data manipulation [Manual]. https://CRAN.R-project.org/package=dplyr

88. Woods, A. J., Antal, A., Bikson, M., Boggio, P. S., Brunoni, A. R., Celnik, P., Cohen, L. G., Fregni, F., Herrmann, C. S., Kappenman, E. S., Knotkova, H., Liebetanz, D., Miniussi, C., Miranda, P. C., Paulus, W., Priori, A., Reato, D., Stagg, C., Wenderoth, N., & Nitsche, M. A. (2016). A technical guide to tDCS, and related non-invasive brain stimulation tools. Clinical Neurophysiology: Official Journal of the International Federation of Clinical Neurophysiology, 127(2), 1031–1048. 10/f77qbf

89. Wurzman, R., Hamilton, R. H., Pascual-Leone, A., & Fox, M. D. (2016). An Open Letter Concerning Do-It-Yourself Users of Transcranial Direct Current Stimulation. Annals of Neurology, 80(1), 1–4. 10/gf935w

90. Zettler, P. J. (2017). tDCS Research in a World With FDA Regulation. AJOB Neuroscience, 8(1), 1–2. 10.1080/21507740.2017.1293192

